# Unraveling HCV Diversity and Resistance in Viet Nam: Implications for Treatment

**DOI:** 10.64898/2026.03.14.26348368

**Authors:** Chau Le Ngoc, Haiting Chai, George Airey, Tanmay Das, Daisy Jennings, Mike Xu, Barnaby Flower, Mahdi Moradi Marjaneh, Leanne McCabe, Hung Le Manh, Chau Nguyen Van Vinh, Thuan Dang Trong, Thach Pham Ngoc, Huong Vu Thi Thu, Guy E Thwaites, H. Rogier van Doorn, Jeremy Day, Evelyne Kestelyn, Tan Le Van, Motiur Rahman, VIETNARMS Study Group, Sarah Pett, Eleanor Barnes, A. Sarah Walker, Graham S. Cooke, M. Azim Ansari

## Abstract

**Background:** Viet Nam has one of the world’s most diverse hepatitis C virus (HCV) epidemics, dominated by genotype 6. Understanding pre-treatment resistance-associated substitutions (RASs) particularly in under-studied genotype 6 is essential to protect cure rates and guide national elimination strategies. We aimed to evaluate the landscape of viral diversity and baseline drug resistance in Vietnam.

**Methods:** We utilized whole-genome sequencing to analyze HCV isolates from a cohort of 1,649 patients enrolled in six clinical studies in Viet Nam between 2013 and 2023. The study assessed genotype and subtype distribution, associations with demographic and clinical variables, and prevalence of known and putative RASs in NS3, NS5A, and NS5B relevant to DAAs used in Viet Nam.

**Findings:** Phylogenetic analysis revealed that genotype 6 was dominant (50.3%, 829/1,649). We observed distinct geographical and demographic partitioning: genotype 2 was concentrated in the south and associated with older age and HIV co-infection, while genotype 3 was clustered in the north among younger males. Clinically relevant RASs were detected in 37.9% (617/1,630) of patients, with the highest burden in NS5A region. Genotypes 2 and 3 displayed near-universal intrinsic resistance. Among genotype 6 infections, subtype 6a frequently carried L28F mutation (43.3%, 181/418), whereas subtype 6e remained largely susceptible.

**Interpretation:** Viet Nam is characterized by a complex, genotype 6-predominant HCV epidemic with significant reservoirs of natural resistance. The high-level resistance mutations in genotypes 2 and 3 suggests that “pan-genotypic” regimens may face efficacy gaps, highlighting the need for subtype-level molecular surveillance to guide national treatment policies.

## Introduction

Hepatitis C virus infection represents a significant global health challenge, being a primary cause of chronic liver disease, which often progresses to severe outcomes like cirrhosis and hepatocellular carcinoma over decades. The World Health Organization estimated that in 2022, approximately 50 million people lived with chronic HCV globally, with about 1 million new infections each year [1]. The burden varies significantly by region. In Southeast Asia, and specifically Viet Nam, prevalence estimates range from 1% to 4.7% in the general population but can be as high as 89% within high-risk groups, including people who inject drugs (PWIDs), people living with HIV (PLWH), and those frequently receiving blood transfusions or dialysis [2–4].

HCV is an enveloped, positive-sense single-stranded RNA virus of the *Flaviviridae* family. Its genome (∼9.6 kb) encodes a single polyprotein processed into three structural (core, E1, E2) and seven non-structural proteins (p7, NS2, NS3, NS4A, NS4B, NS5A, NS5B). It has been classified into eight genotypes (1-8) and over 100 subtypes, with genotype and subtype influencing disease progression and response to treatment [5]. Globally, genotype 1 (46.2%) and genotype 3 (30.1%) are most common [6]. However, Viet Nam and surrounding Southeast Asian nations exhibit a distinct epidemiological landscape dominated by genotype 6, which accounts for over 50% of infections, particularly in southern and central areas, followed by genotypes 1 and 2 [7, 8].

Transmission primarily occurs via blood-to-blood contact. Injection drug use is a major global driver, alongside historical transmission through unsafe blood transfusions prior to widespread screening. Across resource-limited settings, traditional practices like tattooing or acupuncture with non-sterile tools has also been linked to transmission [4, 9, 10].

Genotype 6, notable for its high genetic diversity with over 30 subtypes, likely originated in Southeast Asia and remains largely concentrated there. In Viet Nam, genotype 6 constitutes 54.4% of cases, mainly in the southern and central regions, with subtypes 6a, 6e, 6h, and 6l being prominent [11]. Phylogenetic studies suggest Viet Nam as a potential source for genotype 6 isolates spreading regionally and globally via migration [2, 7, 8]. Transmission patterns within Viet Nam also show genotype specificity: genotype 1 is strongly linked to PWIDs (up to 70.1% prevalence in this group), whereas genotype 6 is more frequently detected among dialysis patients and those with multiple transfusions, suggesting nosocomial spread [12]. These complex dynamics, influenced by healthcare practices, risk behaviors, and population movement, echo trends seen in neighboring regions [11, 13].

The landscape of HCV treatment has transformed since the discovery of virus in 1989. Early interferon-based therapies have offered limited sustained virologic response (SVR) rates and significant side effects. The advent of direct-acting antivirals, starting with first-generation agents like boceprevir and telaprevir in 2011 have marked a turning point. Second-generation DAAs, such as ledipasvir and sofosbuvir (available by 2013-2014), have enabled highly effective (>90% SVR), shorter, safer, and interferon-free regimens. Viet Nam officially adopted DAAs as first-line treatment in 2016, leading to dramatically improved cure rates, although equitable access remains a challenge [14].

Despite the success of DAAs, the pre-existence or emergence of resistance-associated substitutions (RASs) poses a significant threat to treatment efficacy. RASs are mutations that decrease drug susceptibility, often quantified in vitro by an increased half-maximal effective concentration (EC50). Key examples include substitutions at positions D168 (NS3), Y93 (NS5A), S282, and C316 (NS5B). These substitutions can exist as natural variants within the viral population or emerge under the selective pressure of drug treatment, potentially resulting in treatment failure, particularly in patients with prior DAA exposure, poor adherence, or advanced liver disease [15–17]. The prevalence and clinical impact of RASs vary considerably by HCV genotype. While global analyses suggest genotype 6 has the highest RAS prevalence, detailed data, particularly at the subtype level, is scarce [18]. This knowledge gap hinders the development of optimised treatment strategies for genotype 6, which is critically important in Viet Nam given its prevalence [11, 18].

Therefore, continuous RAS surveillance is essential, particularly in regions like Southeast Asia with high genotype diversity. Existing studies on baseline RASs in treatment-naïve Vietnamese patients are limited and often focus only on a specific region like NS5B [19, 20]. Comprehensive data on the prevalence and patterns of RASs across different genotypes, subtypes, and DAA target regions in Viet Nam are urgently needed. Understanding these patterns is critical for informing clinical practice, guiding retreatment strategies, shaping national HCV management policies, and ultimately achieving HCV elimination goals. This study aimed to understand the prevalence of different HCV genotypes in Viet Nam and their geographical and demographic associations and to characterise the prevalence of pre-existing RASs across a large cohort of diverse HCV isolates collected throughout Viet Nam.

## Materials and Methods

### Study description

Plasma samples were obtained from a total of 1,651 patients confirmed to be positive for Hepatitis C virus. These patients were enrolled in six distinct clinical research studies (01EI, VIZIONS, VHARP001, KAPB, SEARCH-1 and VIETNARMS) conducted across northern, central, and southern Viet Nam between 2013 and 2023. The initial trials from 2013 to 2020 were carried out in Ho Chi Minh city. Enrollment was expanded to include additional sites in Hanoi starting in 2020. Socio-demographic data and results from serological tests were extracted from the respective study databases.

### *HCV RNA* extraction

Viral RNA was extracted from 200μl of plasma using the QIAamp Viral RNA Mini Kit (Qiagen, Germany), with linear acrylamide added as a carrier. RNA was eluted in 30μl of buffer AVE and stored at −80°C until further processing.

### Library preparation and next-generation sequencing (NGS)

Double-stranded cDNA was generated using the NEBNext® Ultra™ II Directional RNA Library Prep Kit for Illumina (New England Biolabs, USA), following manufacturer guidelines with random priming and strand-specific synthesis. Purified cDNA was obtained using SPRIselect magnetic beads (Beckman Coulter, USA). Sequencing libraries were prepared from purified double-stranded cDNA using the NEBNext® Ultra™ II Directional RNA Library Prep Kit and NEBNext® Multiplex Oligos for Illumina® (New England Biolabs, USA), including end repair, adaptor ligation, and PCR enrichment. Final libraries were purified using SPRIselect beads and assessed for fragment size and concentration using the Agilent TapeStation (Agilent Technologies, USA) and Qubit fluorometry (Thermo Fisher Scientific, USA). Target enrichment was performed using the Twist Hybridization and Wash Kit with Amplification Mix (Twist Bioscience, USA) and a custom probe panel, followed by post-capture PCR amplification. Final libraries were quantified, normalized, and sequenced on the Illumina MiSeq platform according to the manufacturer’s instructions.

### Sanger sequencing of samples without NGS genome recovery

Samples that did not yield usable genome sequences by NGS due to insufficient coverage were subjected to targeted Sanger sequencing. Viral RNA was reverse transcribed to cDNA using superscript III reverse transcriptase protocol as described earlier [2], and specific genomic fragments were amplified using sets of primers designed across the HCV genome (Supplementary Table 6). PCR products were purified and sequenced using the Sanger method on an ABI 3130xl DNA Analyzer (Applied Biosystems, USA) according to the manufacturer’s instructions. Resulting chromatograms were inspected for sequence quality, and consensus sequences were generated and aligned to the H77 reference genome (GenBank accession: AY051292.1) to determine genomic positions and confirm subtype assignments.

### Identification of Previously Reported and Novel Resistance-Associated Substitutions

Raw sequencing reads generated on the Illumina MiSeq platform were processed using QuasR (v7.01) to remove low-quality reads (length < 50 bp or median Phred quality < Q30) and cutadapter (v1.7.1) to trim illumine sequencing adapters. Human-derived reads were removed by aligning against the T2T human genome assembly via bowtie2 (v2.2.4). Viral genomes were then assembled using bwa (v0.7.17) and VICUNA (v1.3) based on the refined reads and HCV references from International Committee on Taxonomy of Viruses (ICTV). Consensus nucleotides were called with SHIVER (v1.0.7), requiring at least 50% of aligned reads support at each position. Genome coordinates are reported relative to the H77 reference sequence. For each genomic position, the denominator included only sequences with an amino acid call at that site; sequences containing gaps at the corresponding position were excluded from the calculation.

Following an extensive literature review, previously reported resistance sites associated with DAA treatment outcomes were divided into three categories for the purpose of this study. Our analysis focused on substitutions relevant to the DAAs approved or used in Viet Nam from 2016 to 2024, specifically glecaprevir, grazoprevir, voxilaprevir, daclatasvir, elbasvir, ledipasvir, pibrentasvir, velpatasvir, and sofosbuvir.

### Clinical Resistance-Associated Substitutions (Clinical RASs)

These are amino acid variants at well-characterized NS3, NS5A, and NS5B positions that have been demonstrated *in vitro* or in clinical studies to confer a measurable and clinically meaningful reduction in DAA susceptibility. Their presence is associated with virologic failure, reduced treatment efficacy, or necessitates a regimen adjustment in clinical guidelines (full list in Supplementary Figure 1).

### Sub-clinical Resistance-Associated Substitutions (Sub-clinical substitutions)

These refer to previously reported RASs that are generally not considered clinically significant or are believed to have a minimal impact on drug resistance. These were classified within the context of DAAs used in Viet Nam.

### Putative new RASs

We systematically screened for novel amino acid variants detected at the same genomic positions previously characterized as clinically relevant RAS sites. Specifically, any substitution known to confer resistance in other HCV subtypes but not yet documented in the subtype under investigation was classified as a putative new RAS candidate. These variants represent mutations that may potentially influence DAA susceptibility and were flagged as targets for future phenotypic evaluation to verify their functional impact.

### Data analysis

All statistical analyses were performed using R in RStudio (R Version 4.3.3; RStudio, PBC, Boston, MA, USA).

### Geospatial Analysis

To visualize the geographical distribution of HCV genotypes across Viet Nam, latitude and longitude coordinates were associated with each sample. These data were mapped using administrative boundary shapefiles from the Global Administrative Areas database (GADM) version 4.1.

### Statistics

The study cohort was initially characterized using descriptive statistics, summarizing baseline characteristics such as sex, age at enrollment, viral load, cirrhosis status, and HIV/HBV co-infection, stratified by HCV genotype. Due to non-normality of distributions, age and viral load were compared between genotypes using non-parametric Kruskal-Wallis test, and categorical variables were compared using Chi-squared test.

### Statistical Significance

For all statistical tests, a p-value less than 0.05 was considered statistically significant.

## Results

### Prevalence and Diversity of HCV Genotypes and Subtypes in Viet Nam

Successful whole-genome sequencing was achieved for 1,649 HCV isolates derived from 1,651 collected samples. Among these, 1,285 sequences were generated using next-generation sequencing and 364 were obtained by Sanger sequencing. Phylogenetic analysis indicated that genotype 6 was the most prevalent, identified in 829 sequences (50.3%). This genotype exhibited significant diversity, with subtype 6a being the most dominant (28.4%, 468/1,649), followed by subtype 6e (14.7%, 243/1,649). Notably, subtypes 6a and 6e combined accounted for more than 40% of all sequenced samples. Other subtypes, including 6h, 6l, and 6o, were detected less frequently, while sporadic cases (fewer than 10 samples each) of 6c, 6i, 6k, 6p, 6q, 6t, 6u, and 6xb were also observed.

Genotype 1 was the second most common classification, comprising 649 sequences (39.4%). Within this genotype, subtypes 1a (20.5%, 338/1,649) and 1b (18.9%, 311/1,649) presented in nearly equal proportions. Genotype 2 accounted for 140 sequences (8.5%), primarily represented by subtypes 2a (4.3%, 71/1,649) and 2m (3.6%, 59/1,649). Genotype 3 was the least common, detected in only 31 sequences (1.9%), and consisted of subtypes 3b (1.3%, 21/1,649) and 3a (0.6%, 10/1,649). In summary, four of the eight HCV genotypes were detected in this cohort, with genotypes 6 and 1 predominating (Figure 1).

**Figure 1.**
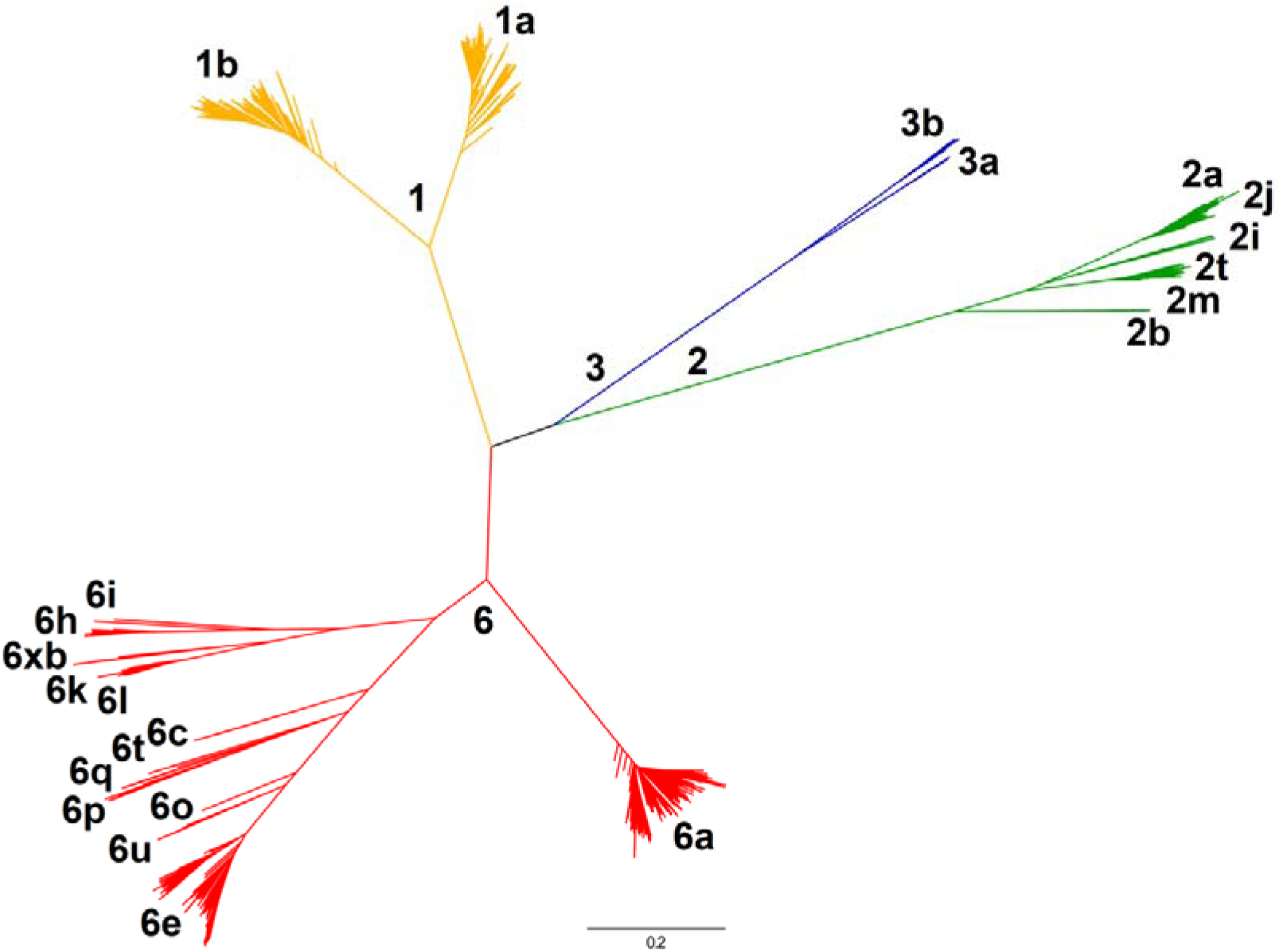
Phylogenetic analysis of 1,649 HCV isolates from Viet Nam. A maximum likelihood tree constructed from whole-genome sequences illustrates the genetic diversity of the cohort. Branches are coloured by genotype: genotype 1 (yellow), genotype 2 (green), genotype 3 (blue), and genotype 6 (red). Major clades corresponding to specific subtypes are labelled at the terminal branches. The scale bar represents 0.2 nucleotide substitutions per site.

### Regional Distribution of HCV Genotypes and Subtypes

Patient enrolment was predominantly conducted in southern Viet Nam (n=1,270), with smaller cohorts contributed by the northern (n=273) and central (n=106) regions (Figure 2). Genotypes 6 and 1, which cumulatively represented 89.7% of the study population, were detected commonly across all three regions (Supplementary Figure 2). Conversely, genotype 2 exhibited a distinct geographical restriction, being identified almost exclusively in southern Viet Nam. In the Mekong River Delta specifically, genotype 2 accounted for approximately 16.3% (80/490) of infections (Supplementary Table 1). Subtype analysis revealed further regional partitioning within genotype 2: subtype 2m predominated in the Mekong River Delta provinces, whereas subtype 2a was prevalent in both the South East and Mekong River Delta regions. Genotype 3 also displayed a marked geographical pattern, with significantly higher prevalence observed in northern Viet Nam compared to the central and southern regions. This northern concentration was primarily driven by subtype 3b, which constituted the majority of genotype 3 infections in the area (Supplementary Table 2).

**Figure 2.**
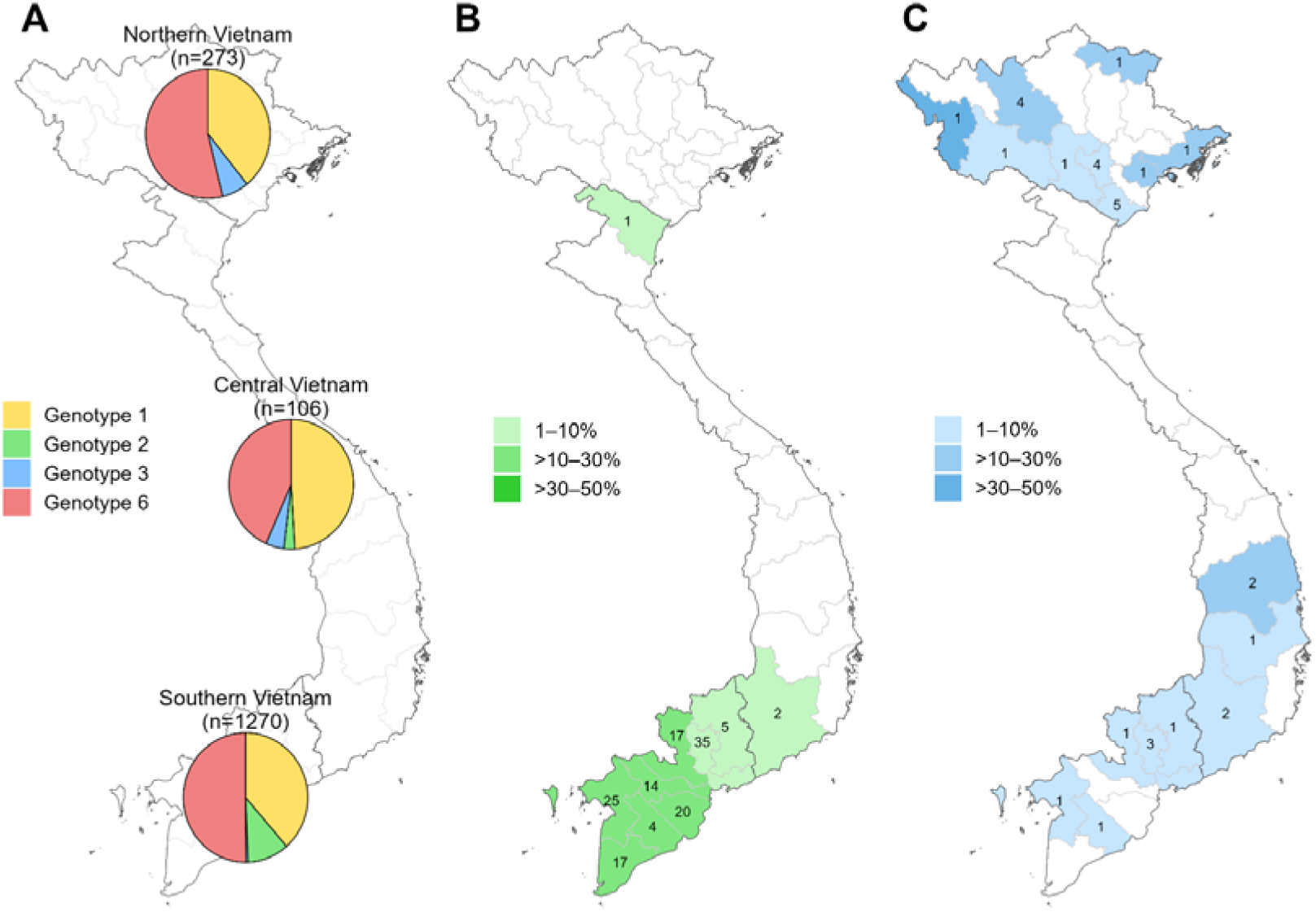
Geographic distribution of HCV genotypes in Viet Nam. (A) Regional distribution of HCV genotypes. Pie charts represent the proportions of genotypes 1 (yellow), 2 (green), 3 (blue), and 6 (red) among sequences obtained from northern (n=273), central (n=106), and southern (n=1,270) Viet Nam. (B–C) Geographical distribution of HCV genotypes 2 (B) and 3 (C) across Viet Nam. Provinces are color-coded based on the prevalence of the specified genotype (light to dark shading indicating proportions of 1–10%, >10–30%, and >30–50%). The numbers displayed on the maps indicate the count of isolates of that specific genotype identified in each province.

### Demographic and Clinical Characteristics of HCV Genotypes in Viet Nam

In this Vietnamese cohort, HCV genotypes showed significant demographic differences. Sex distribution varied considerably between genotypes (p<0.001) with genotype 3 exhibiting strong male predominance (77.4%, 24/31), while genotype 2 had a female majority (59.3%, 83/140). Genotype 1 displayed moderate male predominance (65.2%, 423/649), and genotype 6 presented a nearly balanced profile (54.0% male, 448/829). Age at enrollment also differed significantly across genotypes (p<0.001). Patients with genotype 2 infection were on average older (median age 56 years), those with genotype 3 infection were younger (median age 36 years), while patients with genotypes 1 and 6 infection showed intermediate median ages (42 years and 51 years, respectively). These patterns could potentially reflect different transmission routes or historical exposure periods (Figure 3).

**Figure 3.**
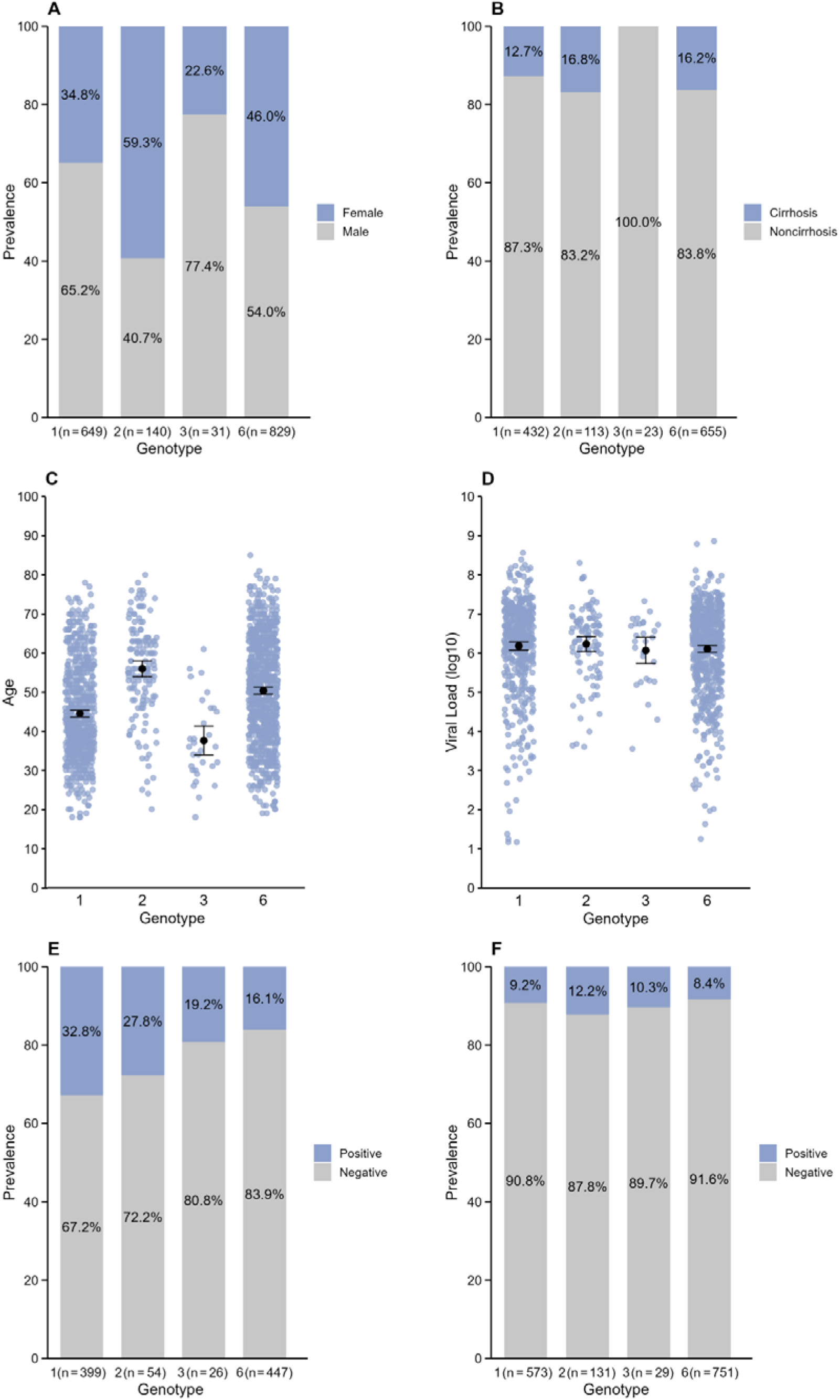
Baseline demographic and clinical characteristics stratified by HCV genotype. (A) Sex distribution across HCV genotypes. (B) Prevalence of cirrhosis by genotype. (C) Age at enrolment by genotype; points represent individual participants. (D) Baseline HCV RNA viral load (loglJlJ IU/mL) by genotype; points represent individual participants. (E–F) Genotype distribution among participants with and without HIV co-infection (E) and HBV co-infection (F); panels are restricted to individuals with available HIV status (n=926) or HBV status (n=1,484). Abbreviations: HBV, hepatitis B virus; HCV, hepatitis C virus; HIV, human immunodeficiency virus; IU, international units.

For a subset of patients, we also collected HIV and HBV co-infection status (926 patients with HIV co-infection status and 1,484 patients with HBV co-infection status). HIV co-infection status showed significant association with HCV genotypes (p<0.001), with genotype 1 more prevalent among PLWH (32.8%, 131/399) and genotype 6 more dominant among HIV-negative individuals (83.9%, 375/447). In contrast, HBV co-infection showed no significant association with HCV genotypes (p=0.506), HBV-positive prevalence was similar across HCV genotypes, observed in 12.2% (16/131) in genotype 2, 10.3% (3/29) in genotype 3, 9.2% in genotype 1 (53/573) and 8.4% in genotype 6 (63/751). Furthermore, baseline viral loads had a similar distribution across genotypes (6.34–6.46 logLJLJ IU/ml, p=0.359). Cirrhosis prevalence varied by genotypes (p=0.056), being highest in genotype 2 infected patients (16.8%, 19/113), followed by genotype 6 (16.2%, 106/655) and genotype 1 (12.7%, 55/432), while no cirrhosis cases were found among the small number of genotype 3 patients (the youngest group, n=23) (Supplementary Table 3).

### Prevalence and Distribution of Resistance

Across this Vietnamese cohort, 37.9% (617/1,630) of sequences contained at least one clinical RAS, with the NS5A protein showing the highest prevalence (26.6%, 422/1,585), followed by NS3 (14.1%, 221/1,565) and with very low prevalence observed for NS5B (1.3%, 21/1,576). This distribution highlighted NS5A as the primary gene for clinically relevant resistance mutations (Figure 4A). Among genotypes, clinical RAS prevalence varied significantly, with genotype 2 sequences exhibiting the highest (83.6%, 117/140), followed by genotype 3 (67.7%, 21/31), genotype 1 (43.0%, 271/630), and genotype 6 (25.1%, 208/829) (Chi-squared p<0.001). These patterns suggest a high baseline risk of resistance in genotype 2 infections. In contrast, genotype 6, the most widely distributed genotype in Viet Nam, demonstrated the lowest overall prevalence of pre-existing clinical RASs (Figure 4B).

**Figure 4.**
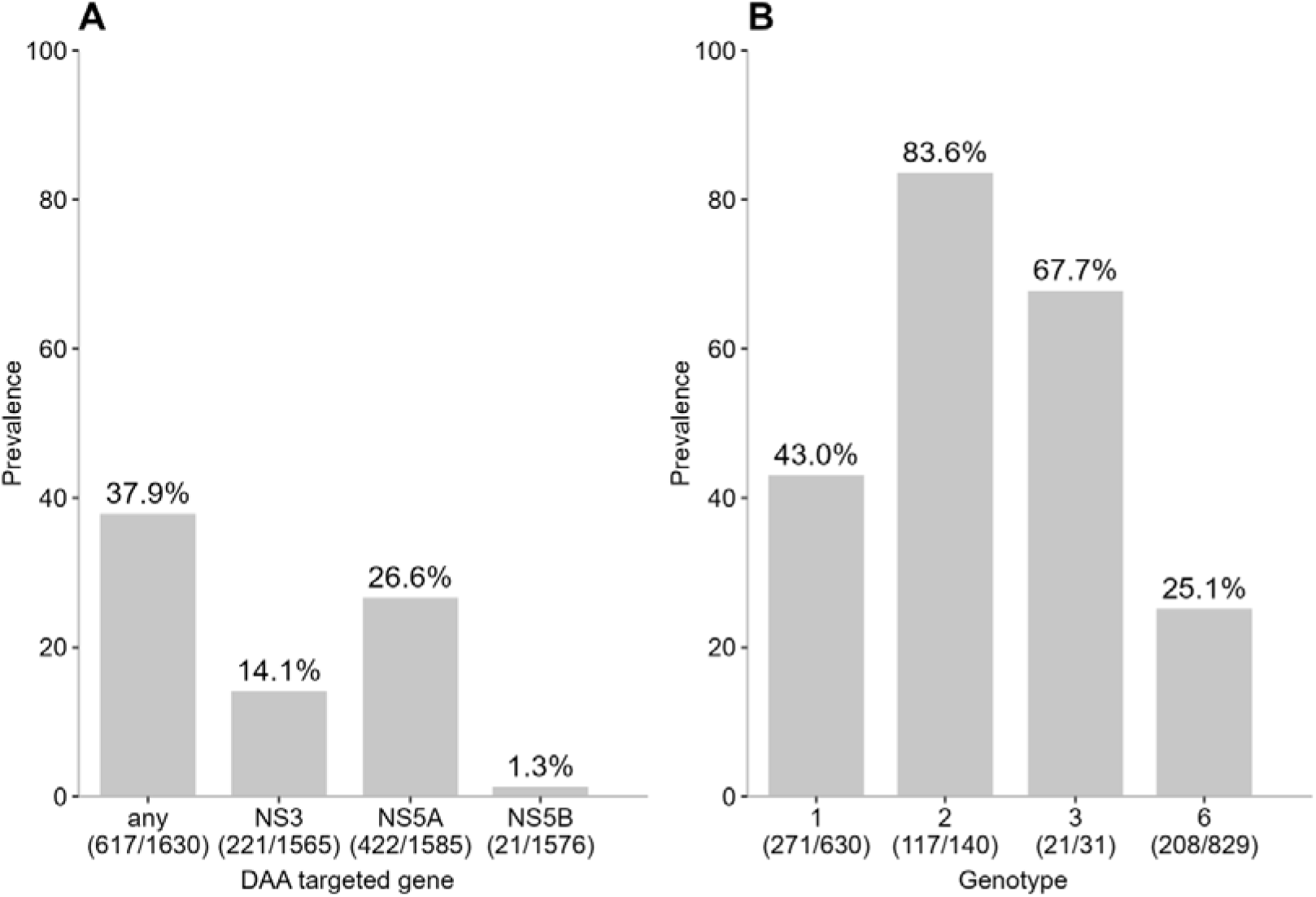
Prevalence of pre-existing clinical RASs in the Vietnamese cohort. (A) Distribution of RASs across the three DAA-targeted viral regions (NS3, NS5A, and NS5B). The “any” category represents the proportion of sequences harboring at least one clinical RAS in any of the three genes. (B) Overall prevalence of RASs stratified by HCV genotype (1, 2, 3, and 6). The percentages above the bars indicate the prevalence of sequences containing at least one RAS, with the corresponding raw counts (number of sequences with RAS / total sequences analyzed) provided below the x-axis labels.

### Genotype 6, subtypes 6a *and* 6e

While data regarding RASs in HCV genotype 6 have historically been limited, our analysis revealed distinct resistance profiles between subtypes 6a and 6e. In subtype 6a, the resistance profile was dominated by substitutions in the NS5A region. The substitution NS5A_L28F was identified as the most prominent clinical RAS, appearing in 43.3% (181/418) of sequences. Other variants at position 28, including L28V (2.4%, 10/418), L28M (0.5%, 2/418), and L28S (0.2%, 1/418), were also detected. Additional known NS5A mutations were observed at lower frequencies, such as L31M (1.4%, 6/417) and T93S (1.9%, 8/428). This pattern of NS5A mutations translated into a high predicted prevalence of resistance to daclatasvir (44.5%, 186/418) and a notably lower resistance profile for velpatasvir (6.2%, 26/418), with minimal predicted resistance to elbasvir (1.4%, 6/418).

Conversely, clinical RAS in the NS3 region of subtype 6a were rare. Substitutions NS3_D168E (0.7%, 3/414) and D168Y (0.2%, 1/414) were detected at minimal frequencies, resulting in very low predicted resistance to NS3 inhibitors such as glecaprevir (0.2%, 1/421) and grazoprevir (0.9%, 4/430). Beyond clinically relevant sites, several sub-clinical RASs were observed in subtype 6a. In the NS3 region, Q41K (0.5%, 2/421) and Q41R (0.2%, 1/421) were noted, while in the NS5A region, sub-clinical variants were identified at position 24 (Q24H), 32 (P32L), 58 (T58A/H/G/S) and position 92 (A92T).

The resistance profile for subtype 6e stood in sharp contrast to 6a. Subtype 6e exhibited an almost complete absence of clinically significant mutations (Figure 5). The only previously clinical reported RAS detected was NS5A_L31M, which was found in a single sample (0.5%, 1/220). Consequently, the predicted resistance to daclatasvir in subtype 6e was negligible.

**Figure 5.**
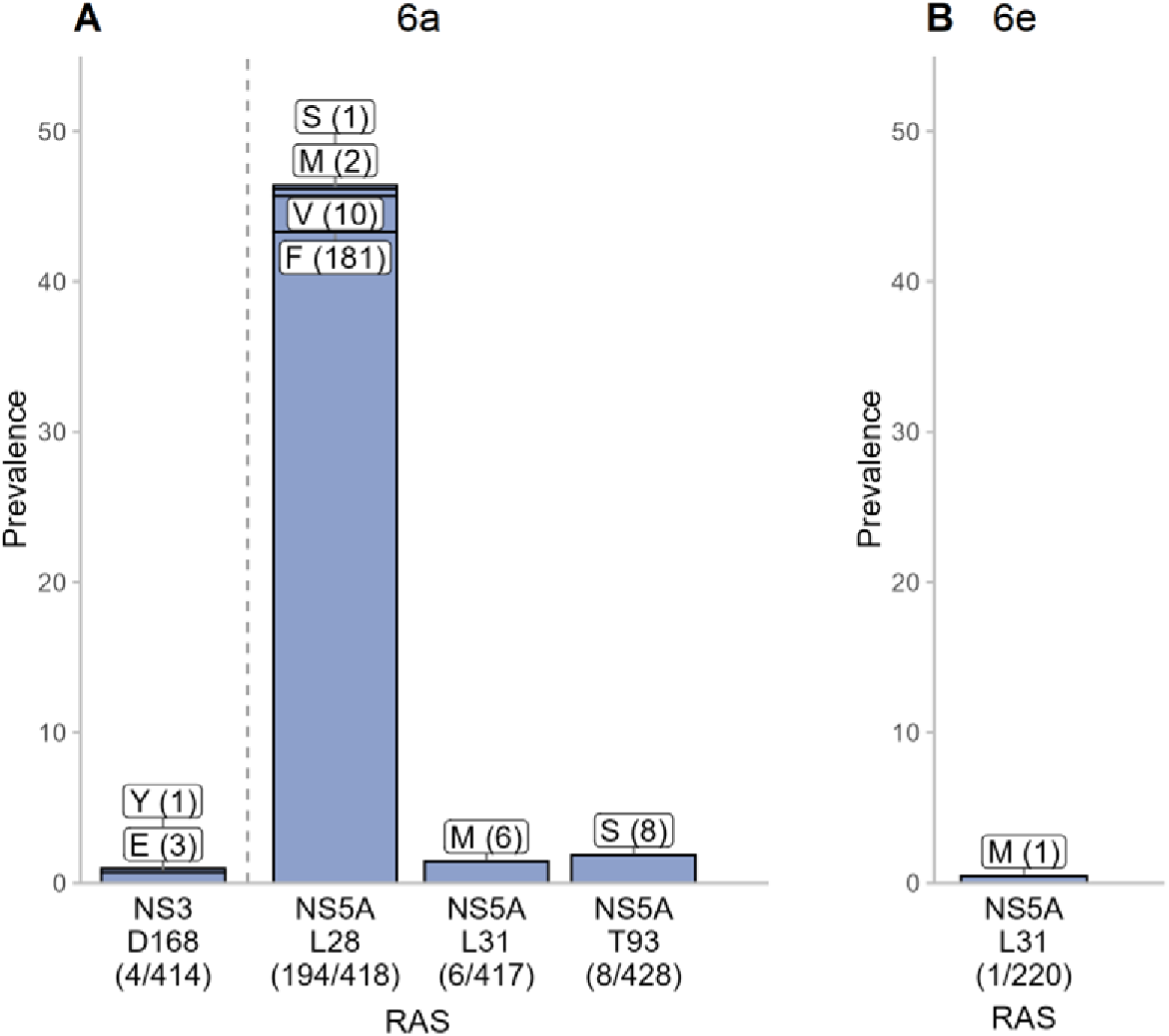
Prevalence and distribution of clinical RASs in HCV subtypes 6a (A) and 6e (B). Bar charts illustrate the prevalence of resistance-associated substitutions at specific polymorphic sites within the NS3, NS5A, and NS5B regions. The x-axis labels identify the specific gene and amino acid position, along with the total count and proportion of samples harboring a mutation. The stacked segments within each bar denote the specific amino acid variants detected; labels indicate the amino acid (single-letter code) and the number of isolates carrying that specific substitution (in parentheses). The dashed vertical line separates RASs located in different viral proteins.

### Subtype 1a

In subtype 1a, the resistance landscape was dominated by the NS3 region. The most common clinical RAS identified was NS3_Q80K, detected in 54.8% (170/310) of sequences. This substitution is associated with substantial resistance to multiple NS3 protease inhibitors, including grazoprevir and voxilaprevir.

In contrast, clinical RASs in the NS5A and NS5B regions were significantly less frequent. In the NS5A region, the most common variation occurred at position 93 (Y93C/H/N/S/T), which was observed in 7.2% (22/304) of samples (Figure 6). Substitutions at this position are associated with reduced susceptibility to a broad range of NS5A inhibitors, including daclatasvir, ledipasvir, pibrentasvir, elbasvir, and velpatasvir. Resistance-associated mutations in the NS5B region were rare, resulting in a very low prevalence of predicted resistance to sofosbuvir (1.3%, 4/316) (Supplementary Table 4).

**Figure 6.**
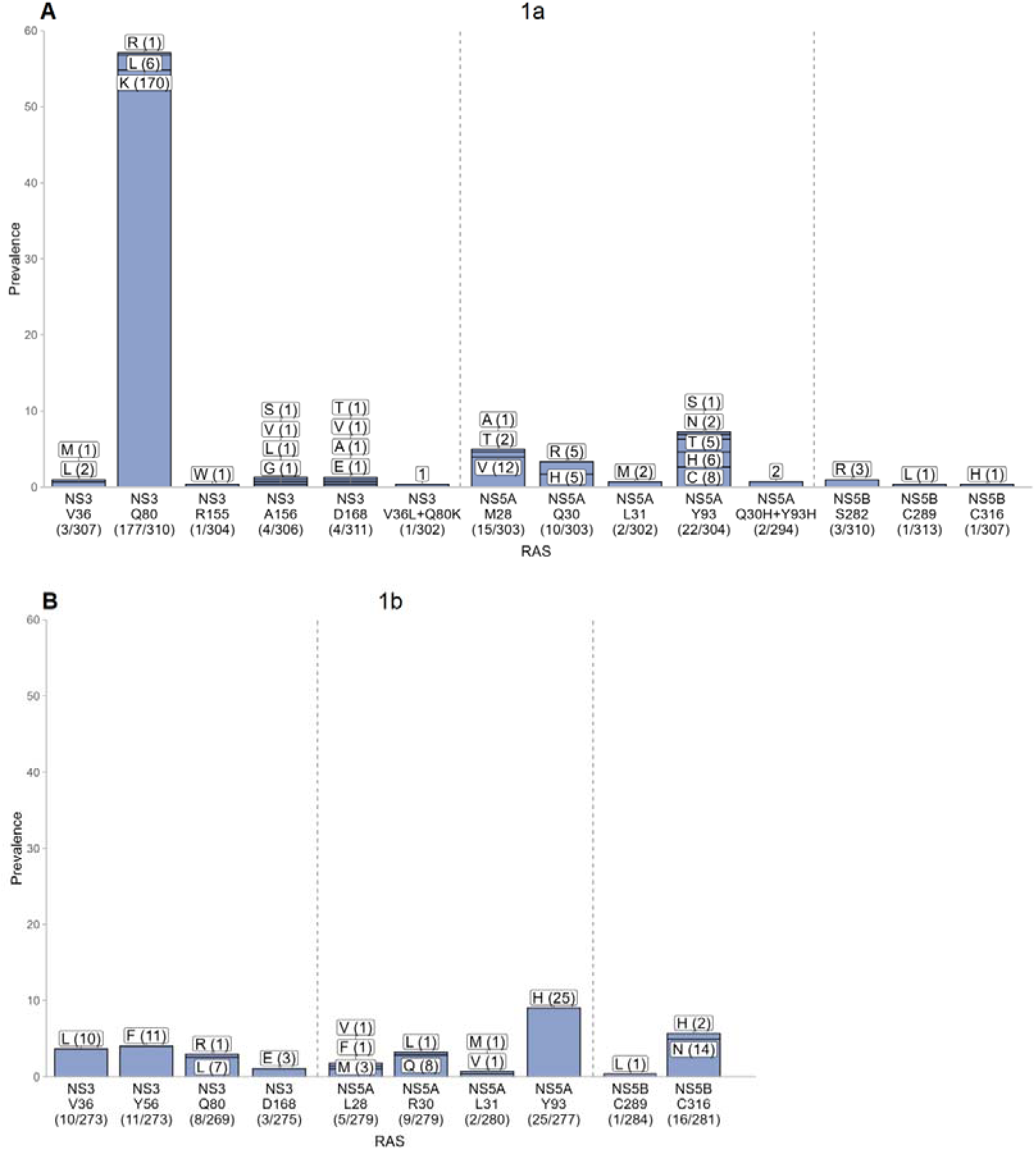
Prevalence and distribution of clinical RASs in HCV subtypes 1a (A) and 1b (B). Bar charts illustrate the prevalence of resistance-associated substitutions at known polymorphic sites within the NS3, NS5A, and NS5B regions. The x-axis labels identify the specific gene and amino acid position, along with the total count and proportion of samples harboring a mutation. The stacked segments within each bar denote the specific amino acid variants detected; labels indicate the amino acid (single-letter code) and the number of isolates carrying that specific substitution (in parentheses). The dashed vertical line separates RASs located in different viral proteins.

Regarding sub-clinical substitutions, most presented at low prevalence (typically ≤1%). However, a few specific polymorphisms were more notable, including NS3_I170V (6.1%, 19/312), NS5A_H58P (5.1%, 16/311), and NS5B_E237G (10.7%, 34/317) (Supplementary Figure 3). These variants have been linked to reduced susceptibility to grazoprevir, daclatasvir, and sofosbuvir, respectively (Supplementary Table 5).

### Subtype 1b

The resistance profile of subtype 1b differed notably from that of subtype 1a, most significantly by the complete absence of the NS3_Q80K substitution, which was highly prevalent in subtype 1a. Instead, the most common clinical RAS identified in subtype 1b was NS5A_Y93H, detected in 9.0% (25/277) of sequences (Figure 6). This substitution is a major contributor to resistance against NS5A inhibitors, conferring reduced susceptibility to daclatasvir, elbasvir, ledipasvir, pibrentasvir, and velpatasvir.

Predicted resistance in subtype 1b was more broadly distributed across DAA classes compared to other subtypes. Measurable resistance frequencies were observed for the NS3 inhibitor grazoprevir (11.5%, 32/279), various NS5A inhibitors (ranging from 9.3% to 13.6%), and the NS5B inhibitor sofosbuvir (5.9%, 17/288) (Supplementary Table 4).

Regarding sub-clinical substitutions, most appeared at low prevalence, typically affecting only a small number of patients. A notable exception was the NS5A_F37L substitution, which was observed at a relatively high frequency (18.1%, 51/282) (Supplementary Figure 3). While currently classified as sub-clinical, the high prevalence of this polymorphism suggests it may warrant further investigation.

### Genotype 2, subtypes 2a *and* 2m

Genotype 2 subtypes exhibited a resistance profile characterized by a remarkably high prevalence of clinical RASs, which were concentrated almost exclusively in the NS5A region. The near-universal presence of specific substitutions across these subtypes strongly suggests that these are not acquired resistance mutations selected by prior treatment pressure, but rather represent the wild-type amino acid consensus (subtype-defining polymorphisms) for these lineages in Viet Nam. While these substitutions may be “natural” to the subtype, they function as potent RASs, conferring inherent resistance that could significantly limit the efficacy of NS5A inhibitor-based regimens.

In subtype 2a, NS5A_L31M and NS5A_Y93N appeared as the major resistance determinants, detected in 90.8% (59/65) and 42.4% (28/66) of sequences, respectively. This intrinsic genetic profile translated into extremely high predicted resistance rates for key NS5A inhibitors: 93.8% (61/65) for daclatasvir, 90.8% (59/65) for elbasvir, and 95.4% (62/65) for velpatasvir. Additionally, the sub-clinical RAS NS5A_T24A was observed in 6.2% (4/65) of subtype 2a sequences. While its individual frequency was low, this pattern warrants attention as it may further contribute to reduced DAA susceptibility.

Subtype 2m displayed a similarly restricted resistance pattern dominated by NS5A_L31M, which was fixed in 98.1% (53/54) of sequences. This naturally occurring polymorphism drove a predicted resistance rate of 98.1% to daclatasvir. Notably, no clinical RAS was detected in the NS3 or NS5B regions for either genotype 2 subtype, confirming that the inherent resistance in this group is NS5A-centric (Figure 7).

**Figure 7.**
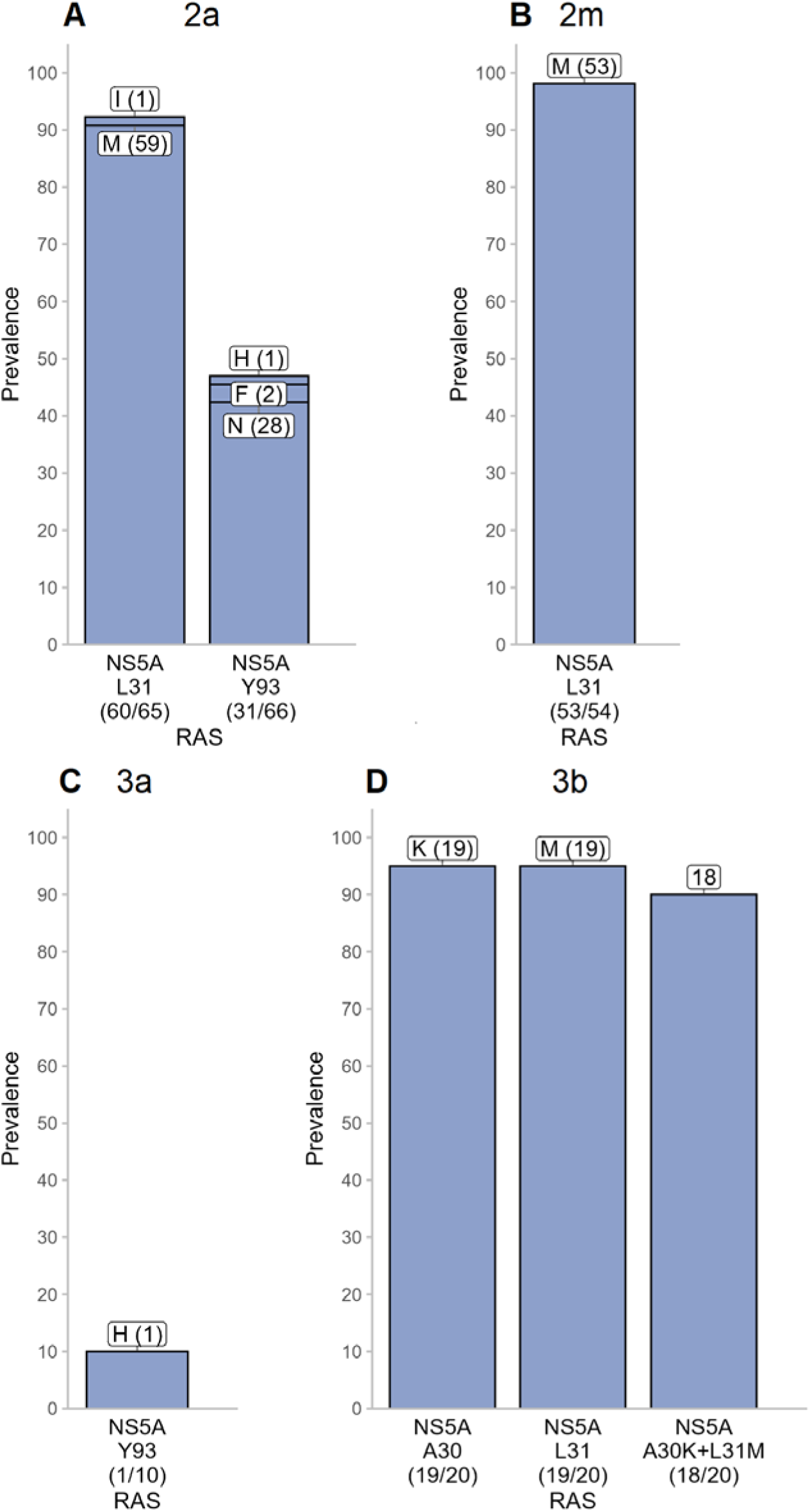
Prevalence and distribution of clinical RASs in HCV subtypes 2a (A), 2m (B), 3a (C), and 3b (D). Bar charts illustrate the prevalence of resistance-associated substitutions at known polymorphic sites within the NS3, NS5A, and NS5B regions. The x-axis labels identify the specific gene and amino acid position, along with the total count and proportion of samples harboring a mutation. The stacked segments within each bar denote the specific amino acid variants detected; labels indicate the amino acid (single-letter code) and the number of isolates carrying that specific substitution (in parentheses). The dashed vertical line separates RASs located in different viral proteins.

### Genotype 3, subtypes 3a *and* 3b

Genotype 3 subtypes displayed strikingly contrasting resistance profiles, highlighting the critical importance of subtype-level discrimination. In subtype 3a, clinically relevant resistance was minimal. NS5A_Y93H was the only previously reported significant clinical RAS detected, appearing in just 10% (1/10) of sequences. While some sub-clinical substitutions were observed such as NS3_A166S (30.0%, 3/10), NS5A_A62L (20.0%, 2/10), NS5B_A150V (40.0%, 4/10), and NS5B_K206E (10.0%, 1/10), the overall profile suggested a population that remains largely susceptible to current DAAs.

In stark contrast, subtype 3b demonstrated a highly resistant profile driven by specific NS5A mutations. The substitutions NS5A_A30K and NS5A_L31M were each detected in 95.0% (19/20) of sequences, with the dual combination (A30K+L31M) present in 90.0% (18/20) of sequences. The near-fixation of these substitutions indicates they are likely subtype-defining wild-type polymorphisms rather than acquired mutations. Consequently, this natural genetic barrier resulted in a predicted resistance rate of 100% to velpatasvir for all subtype 3b isolates. This finding suggests that unlike subtype 3a, subtype 3b infections possess inherent resistance that may make them particularly challenging to treat with standard NS5A-based regimens (Figure 7).

### Impact on Direct-Acting Antivirals

An assessment of the impact of clinical RASs on specific DAA classes revealed that NS5A inhibitors faced the highest overall burden of predicted resistance. Among these, resistance was most prevalent for daclatasvir (25.4%, 381/1,500), followed by velpatasvir (11.7%, 179/1,527), elbasvir (9.2%, 141/1,529), ledipasvir (4.5%, 72/1,585), and pibrentasvir (2.8%, 45/1,584).

NS3 protease inhibitors demonstrated moderate overall resistance frequencies, primarily driven by grazoprevir (13.8%, 215/1,563) and voxilaprevir (11.6%, 181/1,557), whereas predicted resistance to glecaprevir was negligible (0.4%, 6/1,514). The NS5B polymerase inhibitor sofosbuvir exhibited the highest barrier to resistance, with reduced susceptibility predicted in only 1.3% (21/1,576) of isolates (Supplementary Figure 4).

These patterns indicate that while sofosbuvir-based regimens are likely to retain high efficacy across the majority of HCV isolates in this cohort, treatment strategies relying on NS5A inhibitors face significant resistance challenges. However, these aggregate figures obscure critical disparities between genotypes; therefore, a subtype-specific analysis is essential for guiding individualized treatment approaches (Figure 8).

**Figure 8.**
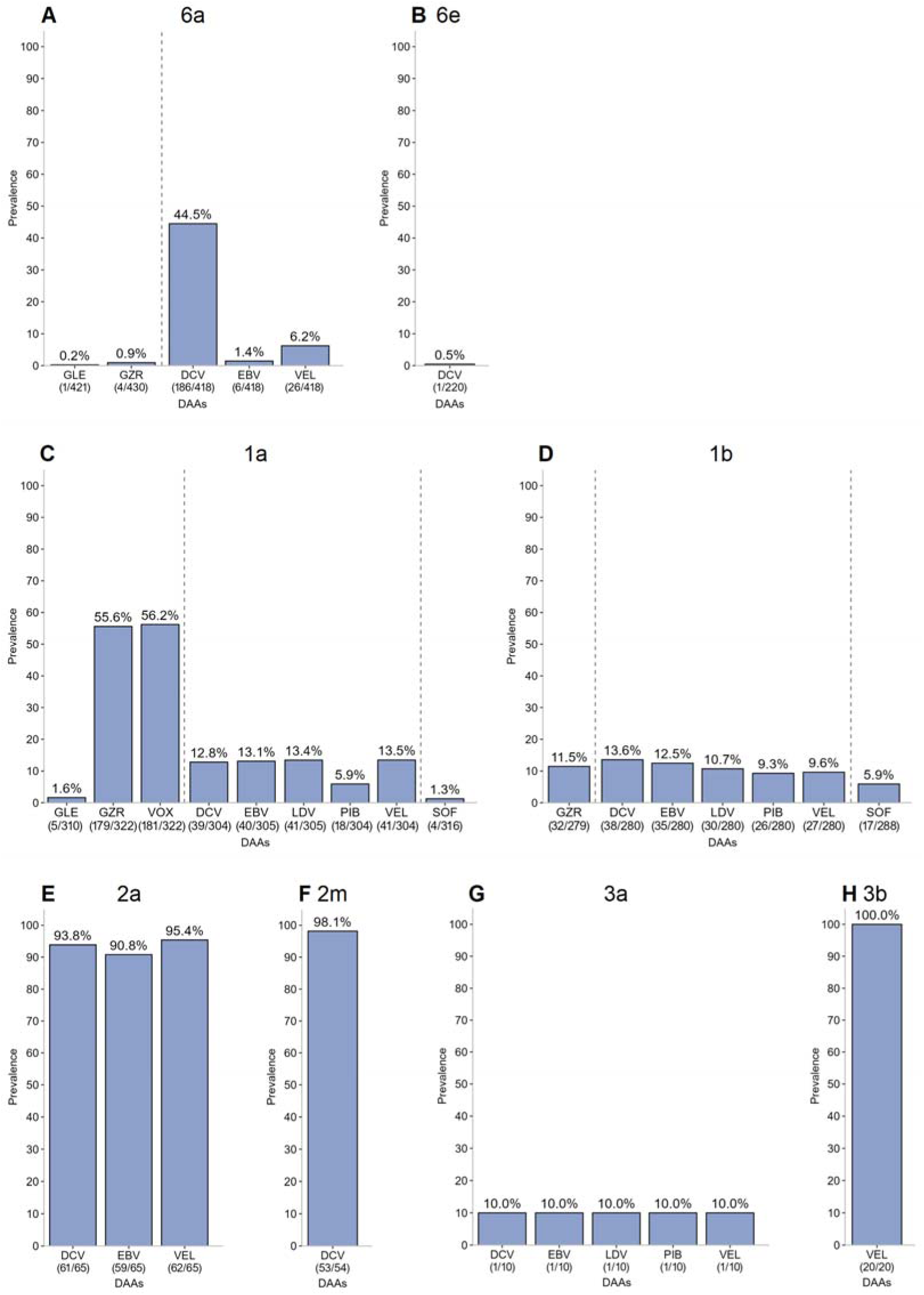
Prevalence of predicted resistance to specific DAAs stratified by HCV subtype. Bar charts illustrate the proportion of isolates harboring at least one clinical RAS associated with resistance to individual direct-acting antivirals. Panels are organized by subtype to highlight distinct resistance profiles. The percentages above the bars indicate the prevalence of predicted resistance, while raw counts (number of resistant sequences / total sequences analyzed) are provided below the x-axis labels. Dashed vertical lines separate DAAs according to their viral protein targets. **Abbreviations:** GLE - glecaprevir; GZR - grazoprevir; VOX - voxilaprevir; DCV - daclatasvir; EBV - elbasvir; LDV - ledipasvir; PIB - pibrentasvir; VEL - velpatasvir; SOF - sofosbuvir.

### Emerging Concerns: Putative Novel RASs

Beyond established resistance mutations, we identified several putative new RAS variants at positions known to confer resistance in other HCV subtypes but not yet characterized in the subtypes under investigation.

In genotypes 2 and 3, high-frequency substitutions were observed in the NS3 and NS5A regions. In subtype 2a, NS3_36L was found in 97.1% (67/69) of sequences and NS5A_30K in 90.8% (59/65). In subtype 2m, the NS3_36L was universally present in 100% (56/56) of sequences, while NS3_56F was detected in 77.6% (45/58) of sequences. Additionally, subtype 2m sequences frequently harboured substitutions at NS5A position 30, specifically 30R (79.6%, 43/54) and 30K (20.4%, 11/54). Similarly, the NS3_36L substitution was fixed (100.0%) in all subtype 3a (10/10) and subtype 3b (20/20) sequences, with subtype 3b also universally carrying NS5A_28M (100.0%, 20/20).

Given that resistance profiles for genotype 6 remain understudied, we identified numerous putative RASs across its diverse subtypes. In subtype 6a, NS3_80K, a clinically significant mutation in genotype 1a, was detected in 96.0% (403/420) of sequences, while NS5B_289L was found in 3.0% (13/433). Subtype 6e exhibited a complex array of novel variants. At NS5A position 28, variants 28V and 28M were found in 57.3% (125/218) and 41.3% (90/218) of subtype 6e sequences, respectively. The substitution NS5A_30S was detected in 97.7% (213/218) of sequences. At position 93, NS5A_93T was dominant (96.0%, 217/226), with 93S appearing less frequently (2.7%, 6/226). Furthermore, NS5B_289L was nearly universal in subtype 6e (99.6%, 225/226).

High-frequency putative RASs were also conserved in other genotype 6 subtypes. In subtype 6h, NS5A_28V was present in 100% (37/37) of sequences, and NS5A_93T in 94.4% (34/36). In subtype 6l, substitutions NS5A_28V (85.0%, 34/40), NS5A_93T (97.5%, 39/40), and NS5B_289L (97.5%, 39/40) were highly prevalent. Finally, subtype 6o displayed novel variants including NS5A_93S (70.0%, 7/10) and NS5B_289L observed in 100% (11/11) of sequences.

The high prevalence and in many instances, fixation of these substitutions strongly suggests that they represent subtype-defining wild-type polymorphisms rather than acquired mutations driven by selective drug pressure. However, this distinction does not diminish their potential clinical significance. Given that these conserved residues occupy critical drug-binding positions established as resistance hotspots in other genotypes, they likely confer a degree of inherent resistance. This phenomenon implies that specific subtypes, particularly within genotype 6, may possess a naturally elevated baseline resistance to current DAA regimens (“natural resistance”). Consequently, although functional validation through phenotypic assays is required to quantify the exact fold-change in susceptibility, the ubiquity of these variants highlights a critical need to include them in molecular surveillance, as they may act as intrinsic barriers to achieving optimal sustained virologic response (Supplementary Figure 5).

## Discussion

This study represents one of the most comprehensive genomic characterizations of Hepatitis C virus in Viet Nam to date. By analyzing whole-genome sequences from a large, geographically diverse cohort, we have provided a high-resolution map of the country’s complex HCV epidemic. Our findings confirm the unique dominance of genotype 6 and reveal a landscape of RASs that could pose significant challenges to current treatment paradigms. Specifically, the identification of subtype-defining polymorphisms that confer inherent resistance to NS5A inhibitors highlights the limitations of a “one-size-fits-all” approach to HCV elimination in Southeast Asia.

Our results confirm that the Vietnamese HCV epidemic is characterized by high genetic diversity, driven by the co-circulation of four major genotypes. Consistent with regional data, genotype 6 remains the dominant strain nationally (50.3%), particularly in the southern and northern regions, while genotype 1 predominates in central Viet Nam; together, genotypes 6 and 1 account for almost 90% of infections. Additionally, genotype 2 was almost exclusively concentrated in the south, and genotype 3 displayed distinct regional clustering in the north. This strong association between regions and HCV subtypes suggests that diverse transmission pathways and historical founder effects play a crucial role in viral spread.

We observed significant demographic partitioning among genotypes that likely reflects distinct transmission dynamics. Genotype 3 was strongly associated with younger males (median age 36), potentially indicating more recent transmission networks, whereas genotype 2 was concentrated in older females (median age 56), suggesting historical exposure cohorts. Furthermore, the high prevalence of genotypes 1 and 2 among people living with HIV could indicate a link between these genotypes and high-risk behaviours, such as injection drug use, which has been well-documented in Viet Nam [12]. These associations are critical for public health targeting; for instance, screening programs targeting younger populations may encounter different viral profiles than those screening older, general populations.

A pivotal finding of this study is the extraordinarily high prevalence of pre-existing clinical RASs in genotypes 2 and 3. We observed that certain clinical RASs in the NS5A region, specifically L31M in genotype 2 and the dual A30K+L31M mutation in subtype 3b, were fixed in nearly 100% of sequences. The ubiquity of these substitutions suggests they are not acquired mutations selected by prior therapy, but rather subtype-defining wild-type polymorphisms.

This “natural resistance” could have profound clinical implications. For subtype 3b, the presence of these polymorphisms resulted in a predicted resistance rate of 100% to velpatasvir, a frontline NS5A inhibitor. Similarly, genotype 2 subtypes showed more than 90% predicted resistance to daclatasvir, elbasvir and velpatasvir. This indicates that patients infected with these subtypes possess an intrinsic genetic barrier to specific DAA drugs, independent of their treatment history. Consequently, empirical treatment with NS5A-based therapies without resistance testing may carry a higher risk of virologic failure in these specific subgroups [18, 35–37].

Given that genotype 6 is largely neglected in global clinical trials, our detailed characterization of its subtypes and their RAS profile fills a critical knowledge gap. While genotype 6 had the lowest overall clinical RAS prevalence (25.1%), this aggregate figure masks significant subtype-specific risks. In subtype 6a, the most common subtype in Viet Nam the NS5A_L28F substitution was dominant, conferring reduced susceptibility to daclatasvir in 43.3% of cases. Conversely, subtype 6e appeared largely susceptible to current DAAs.

Furthermore, we identified numerous “putative new RASs” in genotype 6, such as NS3_80K in subtype 6a; NS5A_28V/M, 30S, 93T and NS5B_289L in subtype 6e, which appear to be natural polymorphisms. While their phenotypic impact remains to be fully characterized, their location at critical drug-binding sites suggests they may influence treatment outcomes. This highlights the large genetic diversity of HCV and therefore HCV resistance and the necessity of looking beyond the well-characterized genotype 1 RASs when managing non-1 genotypes.

The findings of this study have immediate relevance for HCV management in Viet Nam. First, the high burden of NS5A resistance mutations across multiple genotypes (2, 3b, and 6a) suggests that regimens relying heavily on NS5A inhibitors should be used with caution or guided by genotypic data where possible. Second, the low prevalence of clinical RASs in the NS5B region (1.3%) confirms that sofosbuvir remains a robust backbone for therapy in this population.

There are limitations to this study, including the recruitment primarily from clinical sites which may introduce selection bias, and the reliance on in silico prediction for putative RASs. Future research must focus on the phenotypic validation of these novel mutations, particularly within genotype 6, to quantify their exact impact on drug susceptibility.

In conclusion, the HCV epidemic in Viet Nam is defined by significant genetic diversity and a high burden of intrinsic resistance in specific subtypes. As Viet Nam moves towards HCV elimination, integrating subtype-level surveillance and RAS testing into national strategies will be essential to optimize treatment efficacy and prevent the expansion of resistant viral populations.

## Data Availability

Data access will follow the Oxford University Clinical Research Unit (OUCRU) data sharing policy. De-identified individual participant data, along with the trial protocol and statistical analysis plan, will be available following publication upon reasonable request through the OUCRU Data Access Committee, subject to completion of a data access request form and approval in accordance with the OUCRU data sharing policy.

## Acknowledgements

We would like to thank all study participants and their families, the investigators, clinical, laboratory, and research members involved in the 01EI, VIZIONS, VHARP001, KAPB, SEARCH-1 and VIETNARMS studies conducted by the Oxford University Clinical Research Unit and collaborating institutions in Vietnam.

