## Supplementary Tables for "Unraveling HCV Diversity and Resistance in Viet Nam: Implications for Treatment"

| **Genotype** | **Region** | **Sub region** | **n (%)** |
| --- | --- | --- | --- |
| **1** | Northern Vietnam | North Mountains and Midlands | 35 (38%) |
|  |  | Red River Delta | 72 (39.8%) |
|  | Central Vietnam | North Central Coast | 13 (59.1%) |
|  |  | Central Highlands | 39 (46.4%) |
|  | Southern Vietnam | South East | 366 (46.9%) |
|  |  | Mekong River Delta | 124 (25.3%) |
| **2** | Central Vietnam | North Central Coast | 1 (4.5%) |
|  |  | Central Highlands | 2 (2.4%) |
|  | Southern Vietnam | South East | 57 (7.3%) |
|  |  | Mekong River Delta | 80 (16.3%) |
| **3** | Northern Vietnam | North Mountains and Midlands | 8 (8.7%) |
|  |  | Red River Delta | 11 (6.1%) |
|  | Central Vietnam | Central Highlands | 5 (6%) |
|  | Southern Vietnam | South East | 5 (0.6%) |
|  |  | Mekong River Delta | 2 (0.4%) |
| **6** | Northern Vietnam | North Mountains and Midlands | 49 (53.3%) |
|  |  | Red River Delta | 98 (54.1%) |
|  | Central Vietnam | North Central Coast | 8 (36.4%) |
|  |  | Central Highlands | 38 (45.2%) |
|  | Southern Vietnam | South East | 352 (45.1%) |
|  |  | Mekong River Delta | 284 (58%) |

***Supplementary Table 1.*** *Geographic distribution of HCV genotypes among patients. Values represent the number and proportion of cases within each sub-region.*

| **Genotype** | **Subtype** | **Region** | **Sub region** | **n (%)** |
| --- | --- | --- | --- | --- |
| **1** | **1a** | Northern Vietnam | North Mountains and Midlands | 21 (22.8%) |
|  |  |  | Red River Delta | 37 (20.4%) |
|  |  | Central Vietnam | North Central Coast | 8 (36.4%) |
|  |  |  | Central Highlands | 18 (21.4%) |
|  |  | Southern Vietnam | South East | 194 (24.9%) |
|  |  |  | Mekong River Delta | 60 (12.2%) |
|  | **1b** | Northern Vietnam | North Mountains and Midlands | 14 (15.2%) |
|  |  |  | Red River Delta | 35 (19.3%) |
|  |  | Central Vietnam | North Central Coast | 5 (22.7%) |
|  |  |  | Central Highlands | 21 (25%) |
|  |  | Southern Vietnam | South East | 172 (22.1%) |
|  |  |  | Mekong River Delta | 64 (13.1%) |
| **2** | **2a** | Central Vietnam | North Central Coast | 1 (4.5%) |
|  |  |  | Central Highlands | 1 (1.2%) |
|  |  | Southern Vietnam | South East | 41 (5.3%) |
|  |  |  | Mekong River Delta | 28 (5.7%) |
|  | **2b** | Southern Vietnam | Mekong River Delta | 2 (0.4%) |
|  | **2i** | Central Vietnam | Central Highlands | 1 (1.2%) |
|  |  | Southern Vietnam | South East | 1 (0.1%) |
|  |  |  | Mekong River Delta | 1 (0.2%) |
|  | **2j** | Southern Vietnam | Mekong River Delta | 4 (0.8%) |
|  | **2m** | Southern Vietnam | South East | 15 (1.9%) |
|  |  |  | Mekong River Delta | 44 (9%) |
|  | **2t** | Southern Vietnam | Mekong River Delta | 1 (0.2%) |
| **3** | **3a** | Northern Vietnam | Red River Delta | 4 (2.2%) |
|  |  | Central Vietnam | Central Highlands | 2 (2.4%) |
|  |  | Southern Vietnam | South East | 2 (0.3%) |
|  |  |  | Mekong River Delta | 2 (0.4%) |
|  | **3b** | Northern Vietnam | North Mountains and Midlands | 8 (8.7%) |
|  |  |  | Red River Delta | 7 (3.9%) |
|  |  | Central Vietnam | Central Highlands | 3 (3.6%) |
|  |  | Southern Vietnam | South East | 3 (0.4%) |
| **6** | **6a** | Northern Vietnam | North Mountains and Midlands | 32 (34.8%) |
|  |  |  | Red River Delta | 76 (42%) |
|  |  | Central Vietnam | North Central Coast | 3 (13.6%) |
|  |  |  | Central Highlands | 21 (25%) |
|  |  | Southern Vietnam | South East | 201 (25.8%) |
|  |  |  | Mekong River Delta | 135 (27.6%) |
|  | **6c** | Northern Vietnam | Red River Delta | 1 (0.6%) |
|  | **6e** | Northern Vietnam | North Mountains and Midlands | 7 (7.6%) |
|  |  |  | Red River Delta | 11 (6.1%) |
|  |  | Central Vietnam | North Central Coast | 4 (18.2%) |
|  |  |  | Central Highlands | 7 (8.3%) |
|  |  | Southern Vietnam | South East | 113 (14.5%) |
|  |  |  | Mekong River Delta | 101 (20.6%) |
|  | **6h** | Northern Vietnam | North Mountains and Midlands | 6 (6.5%) |
|  |  |  | Red River Delta | 9 (5%) |
|  |  | Central Vietnam | Central Highlands | 3 (3.6%) |
|  |  | Southern Vietnam | South East | 3 (0.4%) |
|  |  |  | Mekong River Delta | 17 (3.5%) |
|  | **6i** | Central Vietnam | North Central Coast | 1 (4.5%) |
|  | **6k** | Northern Vietnam | North Mountains and Midlands | 2 (2.2%) |
|  | **6l** | Northern Vietnam | North Mountains and Midlands | 2 (2.2%) |
|  |  | Central Vietnam | Central Highlands | 1 (1.2%) |
|  |  | Southern Vietnam | South East | 19 (2.4%) |
|  |  |  | Mekong River Delta | 18 (3.7%) |
|  | **6o** | Central Vietnam | Central Highlands | 1 (1.2%) |
|  |  | Southern Vietnam | South East | 5 (0.6%) |
|  |  |  | Mekong River Delta | 5 (1%) |
|  | **6p** | Southern Vietnam | South East | 2 (0.3%) |
|  |  |  | Mekong River Delta | 4 (0.8%) |
|  | **6q** | Northern Vietnam | Red River Delta | 1 (0.6%) |
|  | **6t** | Central Vietnam | Central Highlands | 2 (2.4%) |
|  |  | Southern Vietnam | South East | 1 (0.1%) |
|  | **6u** | Central Vietnam | Central Highlands | 3 (3.6%) |
|  |  | Southern Vietnam | South East | 5 (0.6%) |
|  |  |  | Mekong River Delta | 1 (0.2%) |
|  | **6xb** | Southern Vietnam | South East | 3 (0.4%) |
|  |  |  | Mekong River Delta | 3 (0.6%) |

***Supplementary Table 2.*** *Geographic distribution of HCV subtypes among patients. Values represent the number and proportion of cases within each sub-region.*

| **Characteristic** | **Total** N = 1,649 | **Genotype 1** N = 649 | **Genotype 2** N = 140 | **Genotype 3** N = 31 | **Genotype 6** N = 829 | **p-value** |
| --- | --- | --- | --- | --- | --- | --- |
| **age, Median [Q1–Q3]** | 48 [38–59] | 42 [36–53] | 56 [49–64] | 36 [30–46] | 51 [40–61] | **3.8e-31*** |
| **gender, n/N (%)** |  |  |  |  |  | **6.8e-09†** |
| Female | 697/1,649 (42.3%) | 226/649 (34.8%) | 83/140 (59.3%) | 7/31 (22.6%) | 381/829 (46.0%) |  |
| Male | 952/1,649 (57.7%) | 423/649 (65.2%) | 57/140 (40.7%) | 24/31 (77.4%) | 448/829 (54.0%) |  |
| **HIV infection, n/N (%)** |  |  |  |  |  | **3.2e-07†** |
| Positive | 223/926 (24.1%) | 131/399 (32.8%) | 15/54 (27.8%) | 5/26 (19.2%) | 72/447 (16.1%) |  |
| Negative | 703/926 (75.9%) | 268/399 (67.2%) | 39/54 (72.2%) | 21/26 (80.8%) | 375/447 (83.9%) |  |
| **HBV infection, n/N (%)** |  |  |  |  |  | 0.506† |
| Positive | 135/1,484 (9.1%) | 53/573 (9.2%) | 16/131 (12.2%) | 3/29 (10.3%) | 63/751 (8.4%) |  |
| Negative | 1349/1,484 (90.9%) | 520/573 (90.8%) | 115/131 (87.8%) | 26/29 (89.7%) | 688/751 (91.6%) |  |
| **Cirrhosis, n/N (%)** |  |  |  |  |  | 0.056† |
| Cirrhosis | 180/1,223 (14.7%) | 55/432 (12.7%) | 19/113 (16.8%) | 0/23 (0.0%) | 106/655 (16.2%) |  |
| Absence of Cirrhosis | 1043/1,223 (85.3%) | 377/432 (87.3%) | 94/113 (83.2%) | 23/23 (100.0%) | 549/655 (83.8%) |  |
| **Log Baseline Viral Load, Median [Q1–Q3]** | 6.37 [5.62–6.90] | 6.42 [5.77–6.96] | 6.46 [5.75–6.82] | 6.35 [5.32–6.75] | 6.34 [5.56–6.90] | 0.359* |
| * Kruskal–Wallis rank sum test; † Pearson’s Chi-squared test | | | | | | |

***Supplementary Table 3.*** *Baseline demographic and clinical characteristics of patients stratified by HCV genotype. Data are presented as median [interquartile range, Q1–Q3] for continuous variables (age and viral load) and as frequency (n/N, %) for categorical variables. The denominator (N) for HIV infection, HBV infection, and cirrhosis status vary due to the availability of clinical data for these specific parameters. P-values indicate the statistical significance of differences across the four genotype groups. * P-values were calculated using the Kruskal–Wallis rank sum test. † P-values were calculated using Pearson’s Chi-squared test.*

| **RAS** | **Protein** | **Resistant to DAA** | **1a** | **1b** | **2a** | **2b** | **2m** | **3a** | **3b** | **6a** | **6e** |
| --- | --- | --- | --- | --- | --- | --- | --- | --- | --- | --- | --- |
| V36L | NS3 | GZR |  | 3.7 (10/273) |  |  |  |  |  |  |  |
| V36L | NS3 | GZR/VOX | 0.7 (2/307) |  |  |  |  |  |  |  |  |
| V36M | NS3 | GLE/GZR | 0.3 (1/307) |  |  |  |  |  |  |  |  |
| Y56F | NS3 | GZR |  | 4.0 (11/273) |  |  |  |  |  |  |  |
| Q80K | NS3 | GZR/VOX | 54.8 (170/310) |  |  |  |  |  |  |  |  |
| Q80L | NS3 | GZR |  | 2.6 (7/269) |  |  |  |  |  |  |  |
| Q80L | NS3 | VOX | 1.9 (6/310) |  |  |  |  |  |  |  |  |
| Q80R | NS3 | GZR | 0.3 (1/310) | 0.4 (1/269) |  |  |  |  |  |  |  |
| R155W | NS3 | VOX | 0.3 (1/304) |  |  |  |  |  |  |  |  |
| A156G | NS3 | GLE/GZR | 0.3 (1/306) |  |  |  |  |  |  |  |  |
| A156L | NS3 | GZR/VOX | 0.3 (1/306) |  |  |  |  |  |  |  |  |
| A156S | NS3 | GZR | 0.3 (1/306) |  |  |  |  |  |  |  |  |
| A156V | NS3 | GLE/GZR/VOX | 0.3 (1/306) |  |  |  |  |  |  |  |  |
| D168A | NS3 | GLE/GZR/VOX | 0.3 (1/311) |  |  |  |  |  |  |  |  |
| D168E | NS3 | GZR | 0.3 (1/311) | 1.1 (3/275) |  |  |  |  |  | 0.7 (3/414) |  |
| D168T | NS3 | GZR/VOX | 0.3 (1/311) |  |  |  |  |  |  |  |  |
| D168V | NS3 | GLE/GZR/VOX | 0.3 (1/311) |  |  |  |  |  |  |  |  |
| D168Y | NS3 | GLE/GZR |  |  |  |  |  |  |  | 0.2 (1/414) |  |
| V36L+Q80K | NS3 | GZR/VOX | 0.3 (1/302) |  |  |  |  |  |  |  |  |
| L28F | NS5A | DCV |  | 0.4 (1/279) |  |  |  |  |  | 43.3 (181/418) |  |
| L28M | NS5A | DCV/EBV/LDV |  | 1.1 (3/279) |  |  |  |  |  |  |  |
| L28M | NS5A | VEL |  |  |  |  |  |  |  | 0.5 (2/418) |  |
| L28S | NS5A | VEL |  |  |  |  |  |  |  | 0.2 (1/418) |  |
| L28V | NS5A | DCV |  | 0.4 (1/279) |  |  |  |  |  |  |  |
| L28V | NS5A | VEL |  |  |  |  |  |  |  | 2.4 (10/418) |  |
| M28A | NS5A | DCV/EBV/LDV/PIB/VEL | 0.3 (1/303) |  |  |  |  |  |  |  |  |
| M28T | NS5A | DCV/EBV/LDV/PIB/VEL | 0.7 (2/303) |  |  |  |  |  |  |  |  |
| M28V | NS5A | DCV/EBV/LDV/VEL | 4.0 (12/303) |  |  |  |  |  |  |  |  |
| A30K | NS5A | VEL |  |  |  |  |  |  | 95.0 (19/20) |  |  |
| Q30H | NS5A | DCV/EBV/LDV/VEL | 1.7 (5/303) |  |  |  |  |  |  |  |  |
| Q30R | NS5A | DCV/EBV/LDV/PIB/VEL | 1.7 (5/303) |  |  |  |  |  |  |  |  |
| R30L | NS5A | DCV |  | 0.4 (1/279) |  |  |  |  |  |  |  |
| R30Q | NS5A | DCV/EBV |  | 2.9 (8/279) |  |  |  |  |  |  |  |
| L31I | NS5A | VEL |  |  | 1.5 (1/65) |  |  |  |  |  |  |
| L31M | NS5A | DCV |  |  |  |  | 98.1 (53/54) |  |  |  | 0.5 (1/220) |
| L31M | NS5A | DCV/EBV/LDV/PIB/VEL | 0.7 (2/302) |  |  |  |  |  |  |  |  |
| L31M | NS5A | DCV/EBV/LDV/VEL |  | 0.4 (1/280) |  |  |  |  |  |  |  |
| L31M | NS5A | DCV/EBV/VEL |  |  | 90.8 (59/65) |  |  |  |  | 1.4 (6/417) |  |
| L31M | NS5A | DCV/VEL |  |  |  | 100.0 (2/2) |  |  |  |  |  |
| L31M | NS5A | VEL |  |  |  |  |  |  | 95.0 (19/20) |  |  |
| L31V | NS5A | DCV/EBV/LDV/PIB/VEL |  | 0.4 (1/280) |  |  |  |  |  |  |  |
| T93S | NS5A | VEL |  |  |  |  |  |  |  | 1.9 (8/428) |  |
| Y93C | NS5A | DCV/EBV/LDV/VEL | 2.6 (8/304) |  |  |  |  |  |  |  |  |
| Y93F | NS5A | VEL |  |  | 3.0 (2/66) |  |  |  |  |  |  |
| Y93H | NS5A | DCV/EBV/LDV/PIB/VEL | 2.0 (6/304) | 9.0 (25/277) |  |  |  | 10.0 (1/10) |  |  |  |
| Y93H | NS5A | DCV/EBV/VEL |  |  | 1.5 (1/66) |  |  |  |  |  |  |
| Y93N | NS5A | DCV/EBV/LDV/PIB/VEL | 0.7 (2/304) |  |  |  |  |  |  |  |  |
| Y93N | NS5A | DCV/VEL |  |  | 42.4 (28/66) |  |  |  |  |  |  |
| Y93S | NS5A | EBV/LDV/VEL | 0.3 (1/304) |  |  |  |  |  |  |  |  |
| Y93T | NS5A | LDV/VEL | 1.6 (5/304) |  |  |  |  |  |  |  |  |
| A30K+L31M | NS5A | VEL |  |  |  |  |  |  | 90.0 (18/20) |  |  |
| Q30H+Y93H | NS5A | DCV/EBV/LDV/PIB/VEL | 0.7 (2/294) |  |  |  |  |  |  |  |  |
| S282R | NS5B | SOF | 1.0 (3/310) |  |  |  |  |  |  |  |  |
| C289L | NS5B | SOF | 0.3 (1/313) | 0.4 (1/284) |  |  |  |  |  |  |  |
| C316H | NS5B | SOF | 0.3 (1/307) | 0.7 (2/281) |  |  |  |  |  |  |  |
| C316N | NS5B | SOF |  | 5.0 (14/281) |  |  |  |  |  |  |  |

***Supplementary Table 4.*** *Prevalence of RASs to DAAs across HCV subtypes. Each value represents the percentage of samples carrying the variant in the indicated HCV subtype within this Vietnamese cohort.* ***Abbreviations:*** *GLE - glecaprevir; GZR - grazoprevir; VOX - voxilaprevir; DCV - daclatasvir; EBV - elbasvir; LDV - ledipasvir; PIB - pibrentasvir; VEL - velpatasvir; SOF - sofosbuvir.*

| **RAS** | **Protein** | **Resistant to DAA** | **1a** | **1b** | **2a** | **2b** | **2i** | **2j** | **3a** | **6a** |
| --- | --- | --- | --- | --- | --- | --- | --- | --- | --- | --- |
| Q41K | NS3 | VOX |  |  |  |  |  |  |  | 0.5 (2/421) |
| Q41R | NS3 | VOX |  |  |  |  |  |  |  | 0.2 (1/421) |
| V55A | NS3 | GZR/VOX | 0.3 (1/309) |  |  |  |  |  |  |  |
| V107I | NS3 | GZR | 0.3 (1/311) | 0.4 (1/278) |  |  |  |  |  |  |
| S122G | NS3 | GZR | 2.0 (6/295) | 3.6 (10/276) |  |  |  |  |  |  |
| S122N | NS3 | GZR | 1.7 (5/295) |  |  |  |  |  |  |  |
| A166S | NS3 | GLE |  |  |  |  |  |  | 30.0 (3/10) |  |
| I170T | NS3 | GZR | 0.3 (1/312) |  |  |  |  |  |  |  |
| I170V | NS3 | GZR | 6.1 (19/312) |  |  |  |  |  |  |  |
| Q80K+I132V | NS3 | GZR | 1.4 (4/296) |  |  |  |  |  |  |  |
| S122T+V170I | NS3 | GZR |  | 0.4 (1/269) |  |  |  |  |  |  |
| Y56F+S122N | NS3 | VOX |  | 0.4 (1/264) |  |  |  |  |  |  |
| Y56F+S122N+V170I | NS3 | VOX |  | 0.4 (1/262) |  |  |  |  |  |  |
| Y56F+V170I | NS3 | GZR |  | 2.3 (6/265) |  |  |  |  |  |  |
| K24R | NS5A | DCV/LDV/PIB/VEL | 1.0 (3/302) |  |  |  |  |  |  |  |
| Q24H | NS5A | DCV |  |  |  |  |  |  |  | 0.2 (1/415) |
| Q24K | NS5A | DCV/EBV/LDV/PIB/VEL |  | 1.8 (5/280) |  |  |  |  |  |  |
| T24A | NS5A | DCV |  |  | 6.2 (4/65) |  |  |  |  |  |
| T24S | NS5A | DCV |  |  | 3.1 (2/65) |  |  |  |  |  |
| P29R | NS5A | PIB | 0.3 (1/303) |  |  |  |  |  |  |  |
| P32L | NS5A | DCV/LDV/VEL |  |  |  |  |  |  |  | 0.2 (1/417) |
| P32S | NS5A | DCV/LDV |  | 1.4 (4/281) |  |  |  |  |  |  |
| F37I | NS5A | DCV |  | 1.4 (4/282) |  |  |  |  |  |  |
| F37L | NS5A | DCV |  | 18.1 (51/282) |  |  |  |  |  |  |
| Y54L | NS5A | DCV |  | 2.8 (8/286) |  |  |  |  |  |  |
| H58D | NS5A | DCV/EBV/LDV/PIB/VEL | 1.0 (3/311) |  |  |  |  |  |  |  |
| H58P | NS5A | DCV | 5.1 (16/311) |  |  |  |  |  |  |  |
| H58R | NS5A | DCV | 1.6 (5/311) |  |  |  |  |  |  |  |
| P58A | NS5A | DCV |  | 0.4 (1/284) |  |  |  |  |  |  |
| P58L | NS5A | DCV |  | 0.7 (2/284) |  |  |  |  |  |  |
| P58S | NS5A | DCV |  | 3.9 (11/284) |  |  |  |  |  |  |
| P58S | NS5A | VEL |  |  |  | 50.0 (1/2) |  |  |  |  |
| T58A | NS5A | DCV |  |  |  |  |  |  |  | 1.4 (6/425) |
| T58G | NS5A | VEL |  |  |  |  |  |  |  | 0.2 (1/425) |
| T58H | NS5A | VEL |  |  |  |  |  |  |  | 0.2 (1/425) |
| T58S | NS5A | DCV |  |  |  |  |  |  |  | 1.9 (8/425) |
| A62L | NS5A | DCV |  |  |  |  |  |  | 20.0 (2/10) |  |
| E62A | NS5A | PIB | 0.3 (1/311) |  |  |  |  |  |  |  |
| E62D | NS5A | DCV |  | 4.2 (12/283) |  |  |  |  |  |  |
| A92T | NS5A | LDV | 0.3 (1/305) | 2.2 (6/277) |  |  |  |  |  |  |
| A92T | NS5A | VEL |  |  |  |  |  |  |  | 0.2 (1/429) |
| C92S | NS5A | DCV/VEL |  |  | 4.5 (3/67) |  |  |  |  |  |
| F37L+Y54H | NS5A | DCV |  | 5.4 (15/280) |  |  |  |  |  |  |
| F37L+Y93H | NS5A | DCV |  | 1.8 (5/273) |  |  |  |  |  |  |
| L31M+Y54H | NS5A | LDV |  | 0.4 (1/278) |  |  |  |  |  |  |
| Q24K+L28M | NS5A | DCV |  | 1.1 (3/278) |  |  |  |  |  |  |
| Q24K+L28M+R30Q | NS5A | DCV |  | 1.1 (3/278) |  |  |  |  |  |  |
| T24S+L31M | NS5A | DCV |  |  |  |  | 66.7 (2/3) |  |  |  |
| T24S+L31M | NS5A | VEL |  |  | 3.1 (2/65) |  |  | 75.0 (3/4) |  |  |
| Y54H+Y93H | NS5A | DCV |  | 1.8 (5/275) |  |  |  |  |  |  |
| A150V | NS5B | SOF |  |  |  |  |  |  | 40.0 (4/10) |  |
| K206E | NS5B | SOF |  |  |  |  |  |  | 10.0 (1/10) |  |
| E237G | NS5B | SOF | 10.7 (34/317) |  |  |  |  |  |  |  |

***Supplementary Table 5.*** *Prevalence of sub-clinical RASs to DAAs across HCV subtypes. Each value represents the percentage of samples carrying the variant in the indicated HCV subtype within this Vietnamese cohort.* ***Abbreviations:*** *GLE - glecaprevir; GZR - grazoprevir; VOX - voxilaprevir; DCV - daclatasvir; EBV - elbasvir; LDV - ledipasvir; PIB - pibrentasvir; VEL - velpatasvir; SOF - sofosbuvir.*

| **Gene** | **Primer name** | **Sequence (5′–3′)** |
| --- | --- | --- |
| 5'UTR-Core | Core-F1 | GGGGCGACACTCCRCCATG |
| 5'UTR-Core | Core-R1 | GCANGASAGCAGDGCCAGNAGG |
| 5'UTR-Core | Core-F2 | CACTCCCCTGTGAGGAACTWCTG |
| 5'UTR-Core | Core-R2 | GAAGATAGARAAARAGCAAC |
| E1 | E1-F1 | GCAACAGGGAAYYTDCCTGG |
| E1 | E1-R1 | GGNGTGAARCAGTABACAGGNCC |
| E1 | E1-F2 | CCYGGTTGCTCYTTYTCTATC |
| E1 | E1-R2 | GGNGCATARTGCCAGCARTABGG |
| E2-p7 | p7-F1 | GCGGNCCDGTVTACTGYTTCAC |
| E2-p7 | p7-R1 | CRTCVCKVCCNCCBCKDRC |
| E2-p7 | p7-R1a | CCACWCNTGCANBDBGGCCTC |
| E2-p7 | p7-F2 | CCTGTDGTSGTVGGNACBACYG |
| E2-p7 | p7-R2 | GVHBANGADRTABBGNADCCNC |
| NS2 | NS2-F1 | CAVGYNGANGCVGCVKTGGAG |
| NS2 | NS2-R1 | CGGCVCCRTGRWRNRYNSYCC |
| NS2 | NS2-F2 | GAGARBCTBRTARTBCTBAAYG |
| NS2 | NS2-R2 | GCNARNVNGGWNYKDGTRRC |
| NS3 | NS3F1-F1 | GGBTGGMRGCTBCTNGCBCC |
| NS3 | NS3F1-R1 | CCVGGRGGNGTGGCBGTBGC |
| NS3 | NS3F1-R1a | GGNGTNGCBGTBGCGARVAC |
| NS3 | NS3F1-F2 | CSATCACNGCNTAYGCYCARC |
| NS3 | NS3F1-R2 | GCYTGGTCNARVACNGTNCC |
| NS3 | NS3F2-F1 | GGCAARTTYCTBGCNGATGG |
| NS3 | NS3F2-R1 | GMRCAYTCYTCCATCTCRTC |
| NS3 | NS3F2-F2 | GRCAYCTBATTTTCTGYCATTC |
| NS3 | NS3F2-F2a | GAAGAARTGYGAYGAGCTSGC |
| NS3 | NS3F2-R2 | GCARTAVGCKGCBARAGCNGC |
| NS4A-NS4B | NS4-F1 | CNAGYACVTGGGTBHTVGTBG |
| NS4A-NS4B | NS4-R1 | GTGNGTNGGKGANACRTGGTTNCC |
| NS4A-NS4B | NS4-F2 | GAKGAGATGGARGAGTGCKC |
| NS4A-NS4B | NS4-R2 | CGAABGCTATNAGYCKRTTCATCCAC |
| NS5A | NS5AF1-F1 | GAYRTNTGGGABTGGATATG |
| NS5A | NS5AF1-R1 | GGHGCNGATABCTGGCTNGC |
| NS5A | NS5AF1-F2 | GGCTBNVNGYYAARCTYNNNHN |
| NS5A | NS5AF1-R2 | GABAGCTGGCTNGCBGADGAGC |
| NS5A | NS5AF2-F1 | GGCARGAGATGGGHRGCAAYATCAC |
| NS5A | NS5AF2-R1 | GAGTCAAARCANCKGGTRTC |
| NS5A | NS5AF2-F2 | GVRTNGAGTCTGAVAMCAARG |
| NS5A | NS5AF2-R2 | CNGGRTANACDATNWVVCGDGC |
| NS5A | NS5AF2-R2a | CYTCRTTCTTNGCCATRATDG |
| NS5B | NS5B-F1 | CCASATCARCTCCGTSTGGCAG |
| NS5B-3'UTR | NS5B-T-R | AAAAAAAAAAAAAAAAAARR |
| NS5B | NS5B-R1a | GGVGCRWANWKGATGATNBWBCC |
| NS5B | NS5B-F2 | GTGTGGGAGGACYTBCTGGAAGAC |
| NS5B | NS5B-R1 | GTGDCKDGCTGYCTCCCAVGC |
| NS5B | NS5B-R2 | GTCATRGCHTCCGTGAADGCTC |

***Supplementary Table 6. Primers used for amplification of HCV genomic regions by Sanger sequencing.*** *Primer names indicate the target genomic region, the fragment number when a region was amplified in multiple overlapping fragments, the primer direction (F, forward; R, reverse), and the PCR round when nested PCR was used (1, outer PCR; 2, nested PCR). Alternative primers designed for the same amplification step are indicated by the suffix “a”. Primer sequences are shown in the 5′–3′ orientation. Degenerate bases follow the IUPAC nucleotide code.*
