## Supplementary Figures for "Unraveling HCV Diversity and Resistance in Viet Nam: Implications for Treatment"

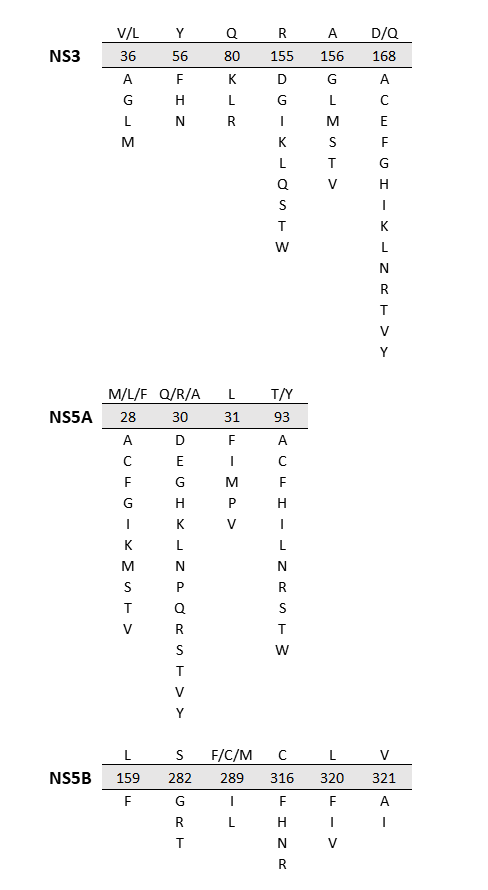

**Supplementary Figure 1.** Reference list of clinically relevant RAS positions in NS3, NS5A, and NS5B genes. This displays key RAS positions within the HCV NS3, NS5A, and NS5B regions. These positions have been well-established in international HCV treatment guidelines or reported in clinical case studies to confer reduced susceptibility to DAAs. This reference was used to identify and classified important RASs in the analysis of Vietnam HCV isolates.

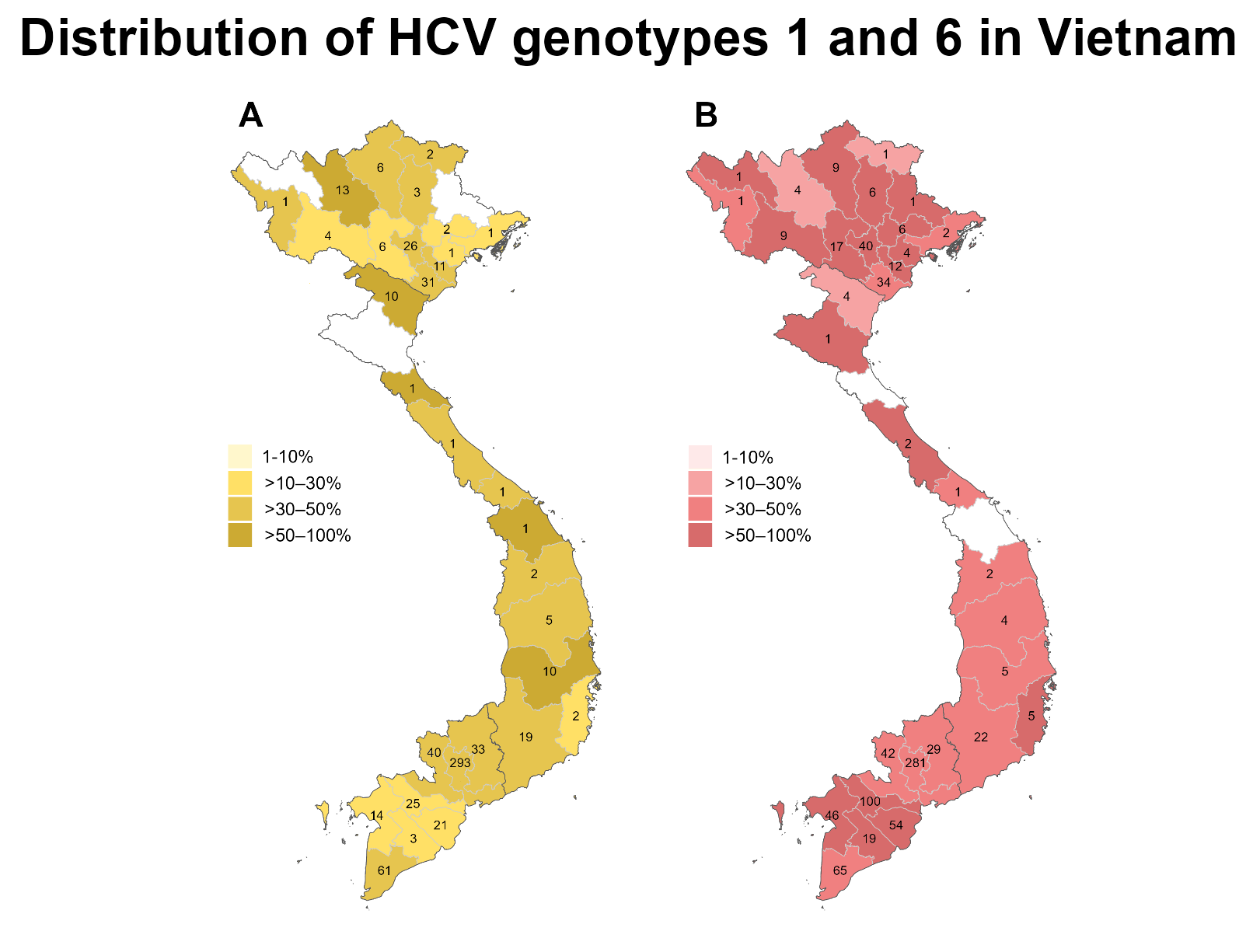

**Supplementary Figure 2.** Geographical distribution of HCV genotypes 1 (A) and 6 (B) across Viet Nam. Each province are shaded according to the number of enrolled patients.

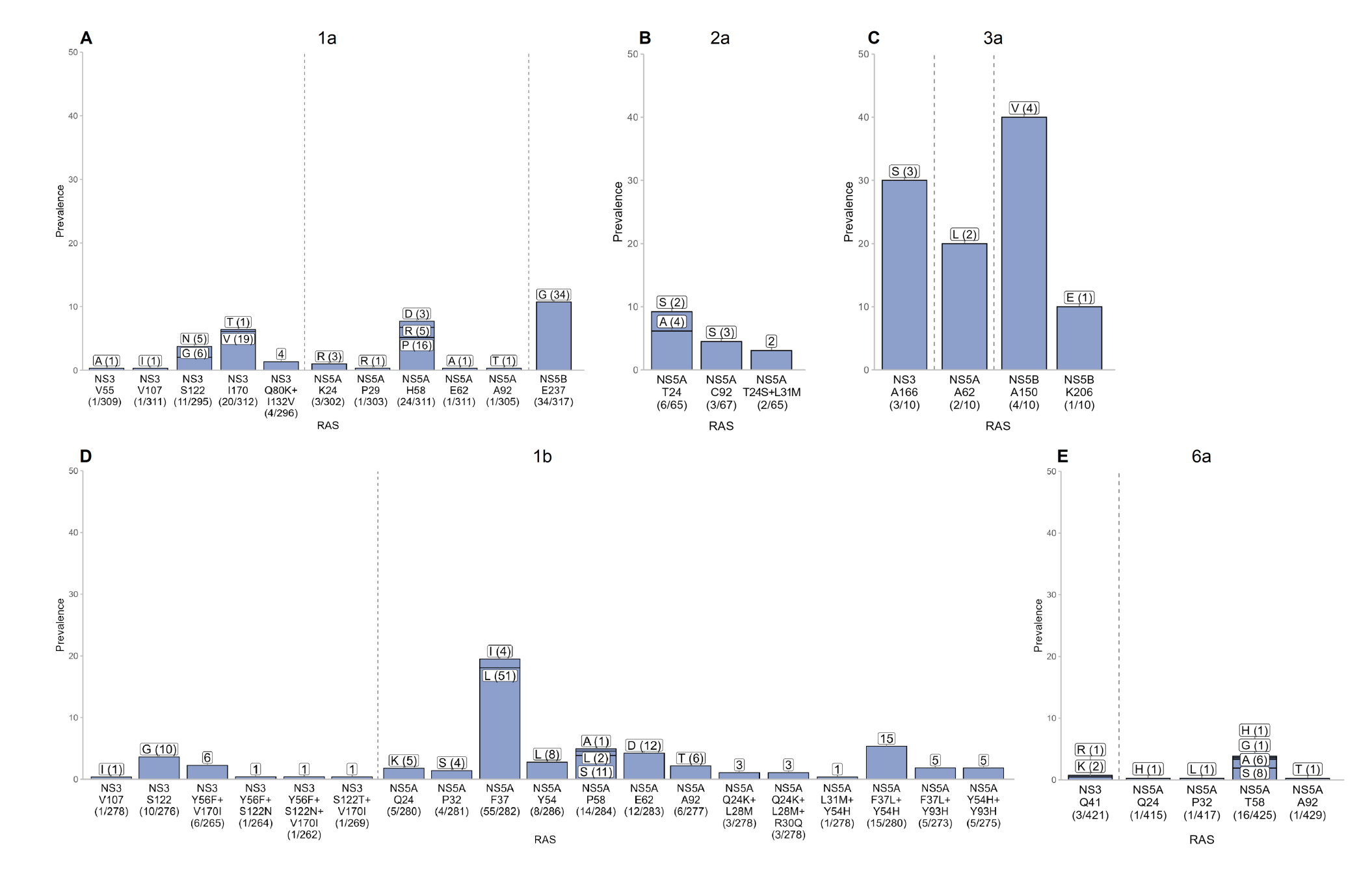

***Supplementary Figure 3.*** *Prevalence of sub-clinical RASs associated with resistance to NS3 protease inhibitors, NS5A inhibitors, and NS5B polymerase inhibitors across HCV subtypes detected in Vietnam. Bar charts show the proportion of sequences harboring mutations at specific amino-acid positions. Labels above the bars indicate the substituted amino acid and the number of isolates carrying that substitution, while counts below the x-axis indicate the number of sequences with the mutation relative to the total sequences analyzed. The dashed vertical line separates RASs located in different viral proteins.*

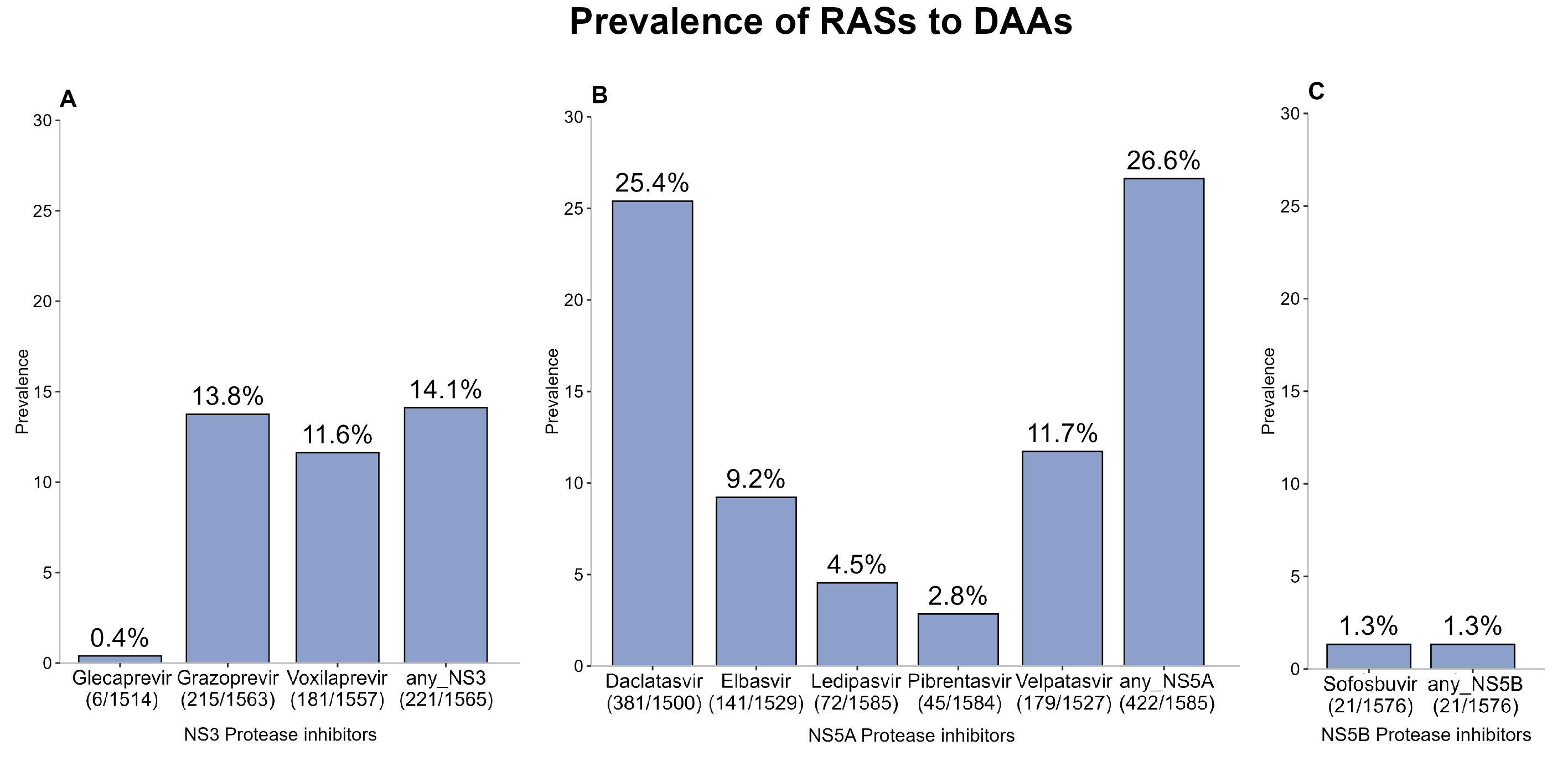

**Supplementary Figure 4.** Prevalence of resistance to DAAs in general Viet Nam HCV cohort. Each bar represents the prevalence of patients whose virus harbored at least one RAS associated with resistance to a specific DAA within the drug class: NS3 protease inhibitors (A), NS5A inhibitors (B) and NS5B polymerase inhibitors (C).

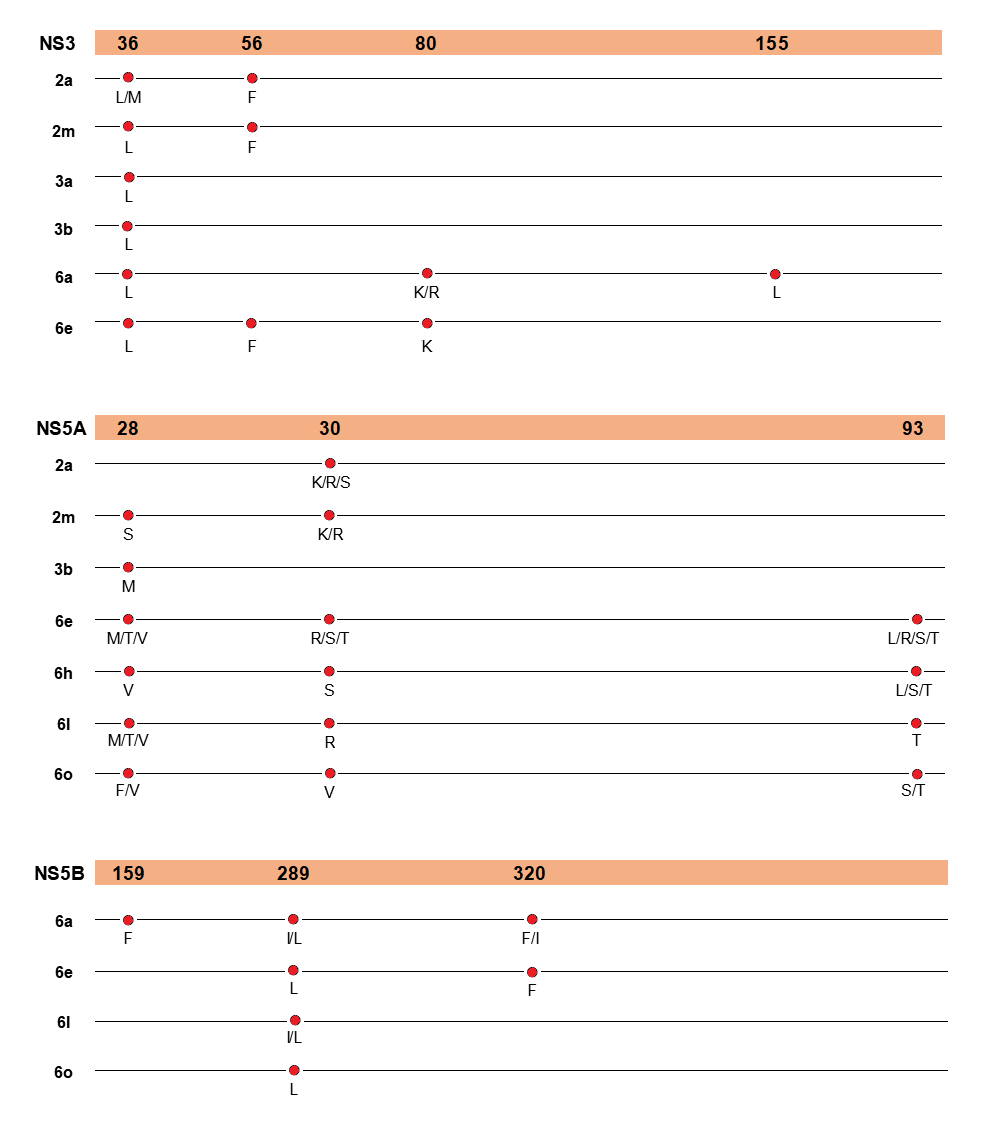

**Supplementary Figure 5.** Schematic representation of putative RASs that were known RASs in other subtypes but newly observed in the examined subtypes in Vietnam isolates. Each horizontal line representes a HCV subtype, with markers showing amino acid substitutions at specific protein positions.
